# Model reduction and analysis: A case study of a malaria control model

**DOI:** 10.1101/2025.08.21.25334195

**Authors:** Maame Akua Korsah, Stuart T. Johnston, Kathryn Tiedje, Karen Day, Camelia R. Walker, Jennifer A. Flegg

## Abstract

Globally rising cases of malaria have prompted concentrated efforts to control malaria transmission, utilising various mathematical models to support the Roll Back Malaria agenda. Many existing models with their specific modifications exhibit rigidity, limiting their application to inform malaria control interventions. This study addresses this limitation by employing a reduction technique on a comprehensive malaria control model to derive a simplified system that preserves the essential dynamics of the original system. We validate the accuracy of the reduced model by comparing the two models via Bayesian MCMC. Based on a simulation study, parameter identifiability analysis and sensitivity analysis, we compare the two models and show that the reduced system exhibits similar transmission characteristics as the full model. Our results demonstrate that the reduced model effectively captures the essential behaviour of the comprehensive model, while providing flexibility and computational efficiency, making it a valuable tool for evaluating and implementing malaria control strategies.

## 1 Introduction

Progress toward global eradication of malaria hangs in the balance, threatened by current rising trends [1, 2, 3]. After several efforts to control malaria transmission, there is still a persistent and increasing disease burden in malaria-endemic areas, resulting from factors such as undetected asymptomatic transmissions, insecticide and drug resistance in mosquitoes and parasites, climate change, and disruptions to healthcare services [4, 5, 6]. Providing a framework to address these setbacks, the WHO Global Technical Strategy for Malaria 2016-2030 aims to reduce malaria incidence and mortality by at least 90% by 2030, with the updated 2024-2030 strategy offering refined approaches for the remaining years based on current trends [7, 8]. Moreover, other initiatives such as the Roll Back Malaria (RBM) partnership and the High-Burden to High-Impact (HBHI) approach target resources toward the highest-burden countries [9, 10]. Although these efforts by stakeholders aim to achieve malaria elimination in endemic areas, a challenge remains in effectively capturing the complex dynamics of malaria transmission and control in mathematical models; which has for some time now played a crucial role in informing decision-making, as well as optimising and projecting the impact of malaria control measures [11, 12, 13, 14, 15, 16, 17].

Malaria transmission models, dating back to the early twentieth century, have evolved from simple compartmental models into complex models in attempts to capture the intrinsic behavioural patterns of the malaria parasites within the mosquito vector and human host, and to study the dynamics of malaria in the presence of socio-economic factors, climate and environmental conditions [11, 12, 18, 19, 20, 21, 22]. Although these models have provided valuable insights, their complexity can limit their flexibility, making it challenging to apply them to real-life data to answer questions related to malaria transmission and control. Building on the Ross-Macdonald model, several comprehensive malaria models have been developed to study the spread and control of the disease [23]. Notably, Manore et al. (2018) developed a comprehensive age-structured model to investigate the impact of intermittent preventive treatment (IPT) on malaria-induced mortality and drug resistance, highlighting the importance of careful IPT implementation [24]. Ochieng (2023) developed a complex model that captures the interactions between human hosts and mosquito vectors amidst seasonal fluctuations and used this model to assess the impact of awareness campaigns on human behaviour in Kenya [25]. Although effective in fitting malaria incidence data (2000-2021), the complexity of the model limits the incorporation of factors such as drug resistance dynamics and climate change [25]. Korsah et al. (2024) also explored a malaria control model considering the transmission dynamics of non-immune and partially immune individuals in the presence of control strategies [26]. This model effectively captures the individual impacts of insecticide-treated nets (ITNs), long-lasting insecticidal nets (LLINs), indoor residual spraying (IRS), and individual measures. However, integrating multiple interventions, drug resistance, and seasonal characteristics as observed in a real-world endemic setting may prove to be a challenge due to the complexity of the model.

Broad applicability in malaria modelling requires a balance between specificity and flexibility in capturing the disease’s fundamental transmission dynamics while accommodating diverse scenarios. However, many current models prioritise narrow questions over broader applicability, sacrificing generality [19]. To enhance malaria control modelling, a more versatile approach is needed, incorporating key factors such as the interactions between humans and the mosquito vector, symptomatic and asymptomatic infections as well as the presence of control strategies. Increasing the complexity of models can lead to over-parameterisation, making it challenging to estimate and identify model parameters accurately [27]. This is particularly problematic when working with limited or noisy data [28]. Thus, striking a balance between model complexity and simplicity is crucial [19]. Model reduction techniques can be employed to simplify complex models in terms of the number of compartments, parameters, or both, while preserving essential dynamics and improving parameter identifiability and estimation. Reduced models in contrast to complex models offer several advantages, such as; reduced computational time (which tends to allow for faster simulations and analyses), improved parameter identifiability and estimation resulting from working with fewer parameters, and increased applicability to different transmission settings and control strategies [29]. Thus, a reduction of model complexity while retaining key model features can potentially expand a model’s effectiveness in fitting different datasets, and increase accuracy in parameter estimation for specific populations, while assessing the impact of the presence of interventions for malaria control strategies.

In this paper, we derive a reduced model to approximate Korsah et al.’s malaria control model (full model for easy reference), analyse the reduction technique, and compare the structural and practical identifiability of the two models. We apply Bayesian Markov Chain Monte Carlo (MCMC) to assess the accuracy of the “reduced model” in capturing the full model’s dynamics and to estimate model parameters. The study is structured as follows; in Section 2 a reduction of the full model is employed and analysed. In Section 3, we assess the accuracy of the reduced model using model-to-model fitting. Finally, we provide a comprehensive discussion of the reduced model in light of the full model and suggest various applications of the reduced model to improve malaria control programs in Section 4.

## 2 Model Reduction and Analysis

### 2.1 A Full Malaria Control Model

In the malaria control model developed by Korsah et al. (2024), the human population is partitioned into six distinct compartments and the mosquito population into three, see Figure 1. The model of Korsah et al. also considered the impact of intervention strategies to provide valuable information to guide decision making in suppressing malaria transmission [26].

**Figure 1.**
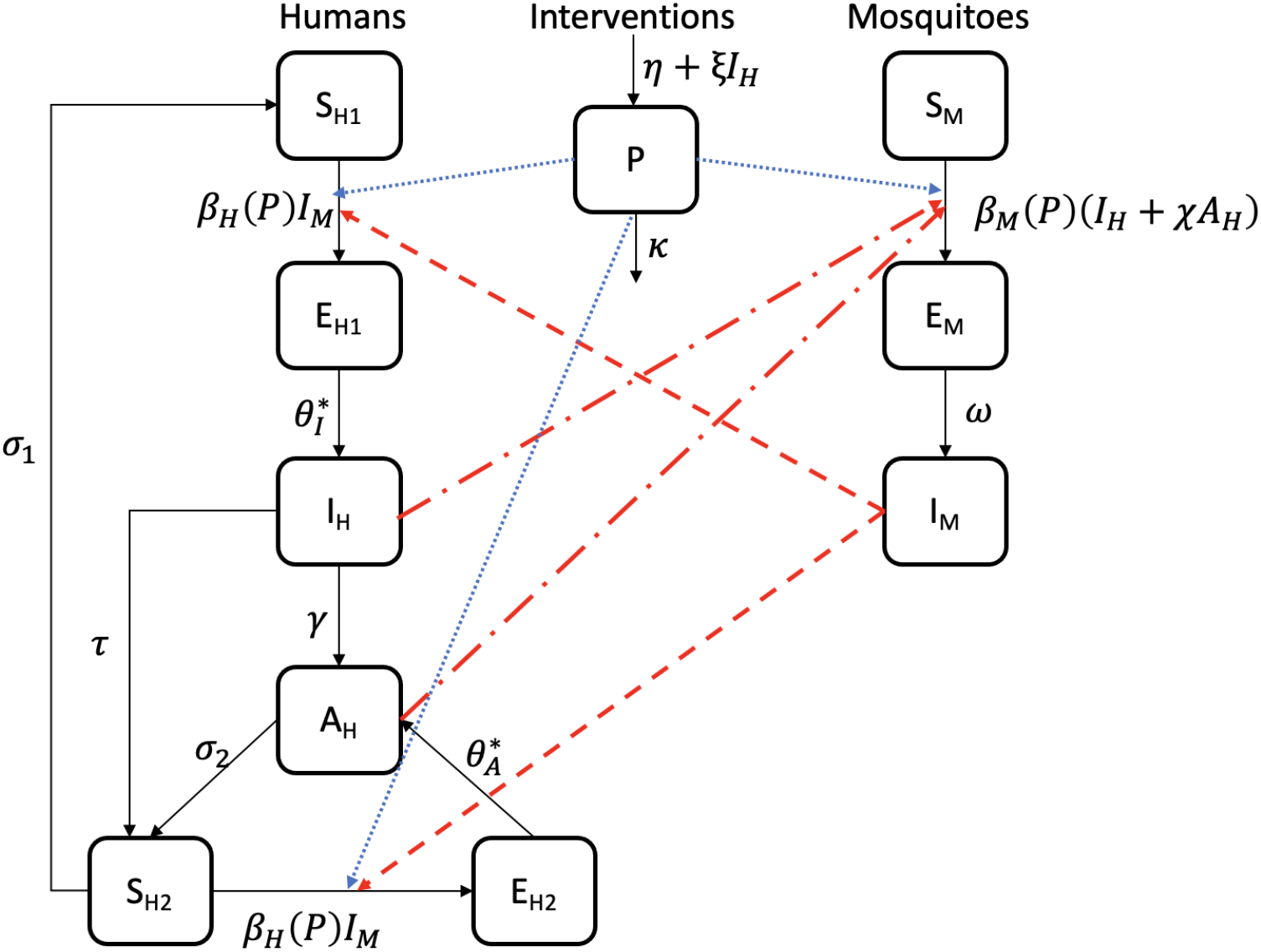
A schematic representation of Korsah et al.’s malaria control model. Transmission from humans to mosquitoes is shown by the red dash-dotted lines, whereas transmission from mosquitoes to humans by the red dashed lines. The blue dotted lines represent the impact of interventions on malaria transmission in both human and mosquito populations. Note that birth and death rates are not shown but are included in the model. Refer to Table 1 for the definitions of the parameters of the model [26].

Note that the parameters *θ*_*A*_ and *θ*_*I*_ in Korsah et al.’s study are redefined as 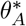 and 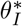 respectively in this study, see Figure 1 and Equation (4) in Appendix A. There are two susceptible and exposed human classes explored in the model, which can be differentiated by the two levels of immunity considered: non-immune and partially immune humans. Non-immune susceptible humans (*S*_*H*1_) can become infected through exposure to malaria parasites, moving them to the first exposed (*E*_*H*1_) class. After a latent phase, they become clinically infectious (*I*_*H*_) and from there, they can either recover into the asymptomatic infectious class *A*_*H*_ or receive treatment into *S*_*H*2_. Partially immune susceptible individuals *S*_*H*2_ can lose their partial immunity status, moving them back into the *S*_*H*1_ class. The full model also accounts for human birth and death rates, infection-induced mortality, and mosquito transmission dynamics [26].

**Table 1:** Definition of the full and reduced model parameters and baseline parameter values employed. For simplicity, we denote “F” and “R” as model references for parameters utilised in the full and reduced models, respectively.

| Parameter | Description | Model ref. | Baseline values | References |
| --- | --- | --- | --- | --- |
| $\Lambda_H$ | influx rate of susceptible humans | F&R | 0.235 persons/day | [34] |
| $\Lambda_M$ | influx rate of susceptible mosquitoes | F&R | 50.5 mosquitoes/day | [34] |
| $\delta$ | mosquito biting rate | F&R | 0.8/ day | [34] |
| $\psi_H$ | transmission success in humans | F&R | 0.5 | [35, 34] |
| $\psi_M$ | transmission success in mosquitoes | F&R | 0.5 | [35, 34] |
| $\mu_H$ | natural death rate of humans | F&R | $4.5 * 10^{-5}$ /day | [35, 34] |
| $\mu_M$ | natural death rate of mosquitoes | F&R | 0.0477 /day | [35, 34] |
| $\tau$ | treatment rate of infected humans | F&R | 0.08 /day | [34] |
| $\omega$ | latency rate in mosquitoes | F&R | 1/9 /day | [36, 37] |
| $\gamma$ | average untreated clinical infection duration rate in humans | F&R | 0.08 /day | [35, 34] |
| $\nu$ | disease induced mortality rate of humans | F&R | 0.08 /day | [35, 34] |
| $\chi$ | relative infectiousness of asymptomatic humans | F&R | 0.8 | - |
| $\eta$ | influx of intervention programs | F&R | 0.3/day | - |
| $\kappa$ | decay rate of intervention programs | F&R | 0.04/day | - |
| $\xi$ | growth rate of programs stimulated by symptomatic infections | F&R | 0.0025/day | - |
| $b$ | constant coefficient of $P$ in the human transmission rate function | F&R | 0.5 | - |
| $c$ | constant coefficient of $P$ in the vector transmission rate function | F&R | 0.5 | - |
| $\theta_I^*$ | latency rate in non-immune humans | F | 1/15 /day | [38] |
| $\theta_A^*$ | latency rate in partially immune humans | F | 1/20 /day | [34] |
| $\sigma_1$ | immunity waning rate of partially immune humans | F | 0.01 /day | [35, 34] |
| $\sigma_2$ | recovery rate from asymptomatic infectiousness | F | 0.01 /day | [35, 34] |
| $\sigma$ | immunity waning rate | R | - | - |
| $\theta_I$ | symptomatic latency rate in humans | R | - | - |
| $\theta_A$ | asymptomatic latency rate in humans | R | - | - |

The full model further incorporates an intervention class (*P*) which represents preventive measures such as vector control measures (IRS, ITNs/LLINs) and IPT, that have a reduction effect on malaria transmission rates. Transmission rate functions in both humans and mosquitoes were defined as:

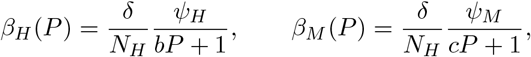

where *b* and *c* are positive-valued constants, *N*_*H*_ is the total human population, *δ* is the biting rate, and *ψ*_*H*_ and *ψ*_*M*_ are the infection probabilities in humans and mosquitoes, respectively [26].

The full model, though comprehensive, is a complex system of 10 dependent variables over time with 21 parameters [26]. The inclusion of additional factors such as seasonality, drug resistance and other real-life data variations would further complicate this model. Hence, we seek a reduced system that incorporates the full model’s essential transmission characteristics such as assessing the impact of control interventions on malaria transmission and accounting for undetectable asymptomatic transmissions from humans to mosquitoes within an endemic population, while being more amenable to analysis and extension.

### 2.2 Reduction of the Full Model - Reduced Model

The full model presented in Figure 1 explicitly captures the differences between partially immune and non-immune people. We now describe a reduced model that combines these two groups, while maintaining dynamics of the remaining compartments. The reduced model divides the human population into four compartments:

- *S*_*H*_ : uninfected individuals susceptible to infection,
- *E*_*H*_ : exposed individuals in the latent phase of infection,
- *I*_*H*_ : clinically infectious humans exhibiting symptoms of infectiousness,
- *A*_*H*_ : asymptomatic infectious individuals with molecular traces of infection.

We reduced the model in this way to address the practical challenge of identifying the number of partially immune and non-immune individuals in a population. This difficulty stems from the lack of variation among serological immunity markers in endemic populations [30, 31]. Thus, the full model 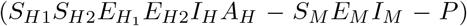 in Equation (4) and Figure 1 condenses to the reduced host-vector system (*S*_*H*_ *E*_*H*_ *I*_*H*_ *A*_*H*_ − *S*_*M*_ *E*_*M*_ *I*_*M*_ − *P*) in Equation (1) and Figure 2 which is an extension of the basic *S*_*H*_ *E*_*H*_ *I*_*H*_ *R*_*H*_ − *S*_*M*_ *E*_*M*_ *I*_*M*_ utilised in previous works [32, 16, 14, 33]. The reduced model captures important features of the full model, including symptomatic transmission, partial immune/asymptomatic transmission, and the effect of intervention strategies.

**Figure 2.**
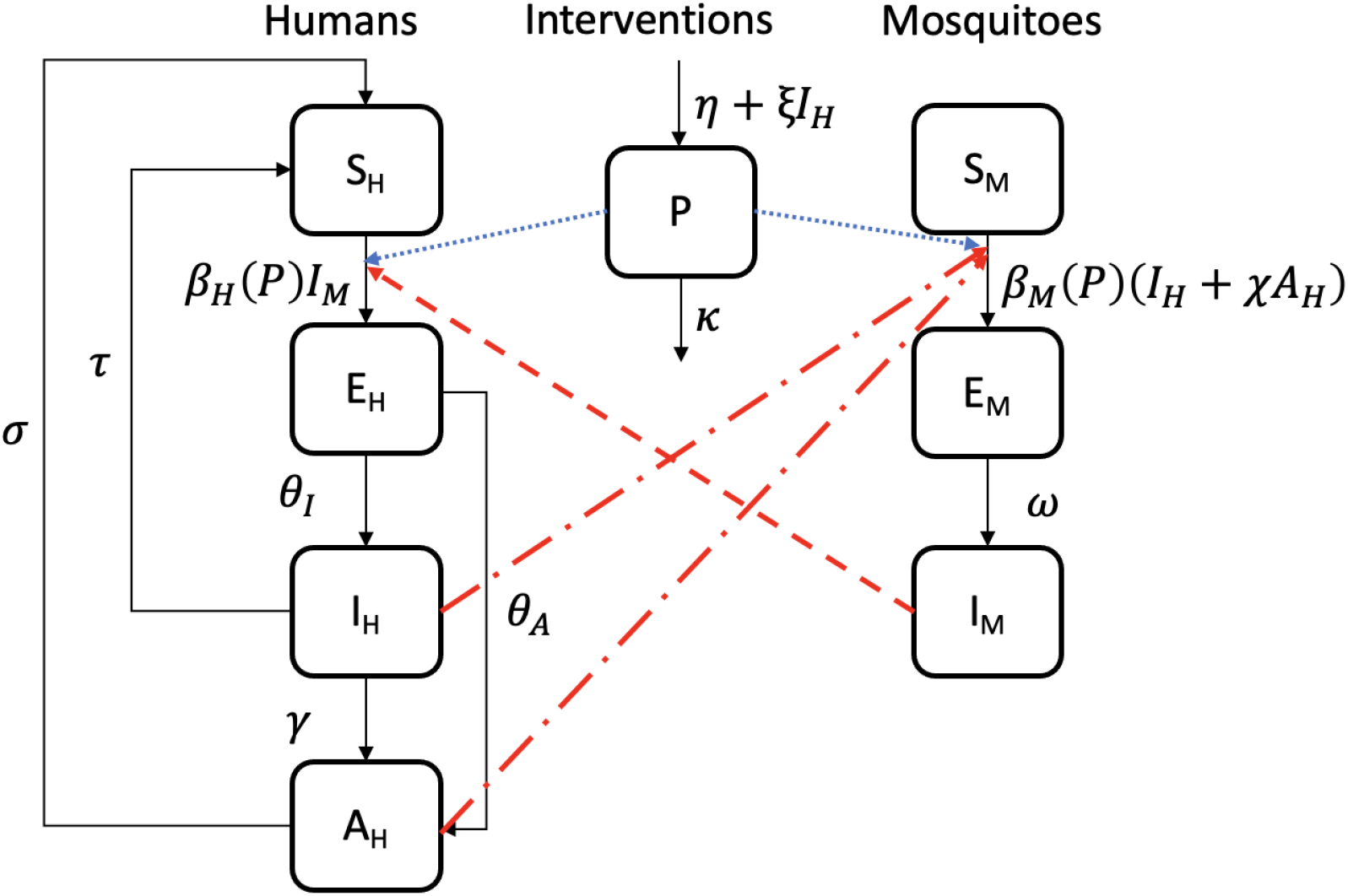
A compartmental diagram of the reduced host-vector model. Transmission from humans to mosquitoes is shown by the red dash-dotted lines, whereas transmission from mosquitoes to humans by the red dashed lines. The blue dotted lines represent the impact of interventions on malaria transmission in both human and mosquito populations. Note that birth and death rates are not shown but are accounted for in the model. Refer to Table 1 for the definitions of the parameters of the model.

The mosquito population and intervention programs are modelled in a similar way to the full model, see Figure 2 and Equation (1):

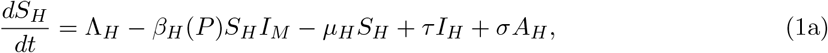

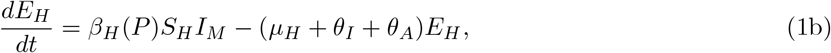

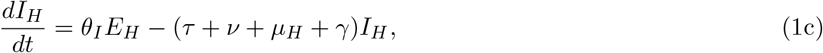

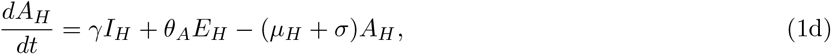

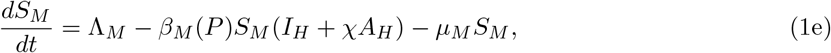

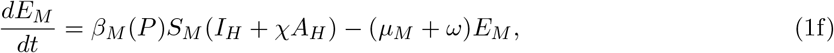

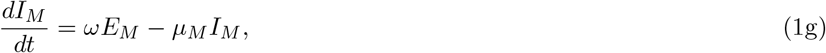

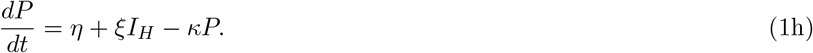

The reduced model has individuals in the susceptible compartment, *S*_*H*_, with population influx rates of Λ_*H*_ (population birth rate), *σ* (immunity waning rate), and *τ* (treatment rate). Individuals in *S*_*H*_ are infected at a rate of *β*_*H*_, moving them into the exposed compartment *E*_*H*_; after which non-immune individuals move into the clinically infectious stage (*I*_*H*_) at a rate of *θ*_*I*_ while partially immune individuals move into the asymptomatic infectious compartment *A*_*H*_ at a rate of *θ*_*A*_. The assumptions of the full model hold for the reduced model, see [26] for details.

### 2.3 Analysis of the Reduction Technique

The alignment of results from the two models depends on how closely each compartment of the models agrees with one another. Thus, we explore the approximation of the full model to the reduced model by equating the dynamics in the susceptible and exposed human compartments. This approach is particularly useful as it allows one to identify and focus on the most influential parameters when comparing complex epidemiological models, thereby reducing computational complexity. Denoting compartments in the full model with superscript “F” and compartments in the reduced model with superscript “R”, we set:

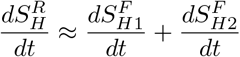

which implies that,

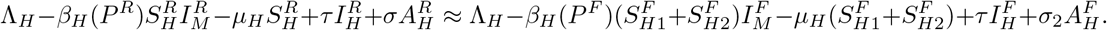

Therefore, the number of susceptible humans in the two models is approximately the same when,

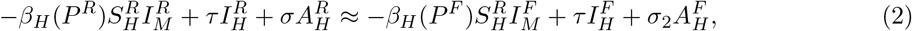

which implies that the disease-driven inflows and outflows of individuals in the susceptible compartments must be approximately equal in both models. This includes the parameters *β*_*H*_ (*P*) (transmission rate function), *τ* (treatment rate), and *σ* (immunity waning rate, which is denoted as *σ*_2_ in the full model), which govern the transmission and progression of the disease within the susceptible human population.

Similarly, by comparing the exposed human compartments of both models;

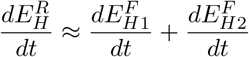

implies that,

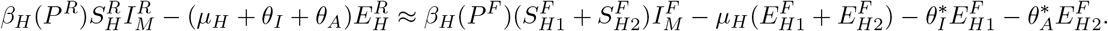

Therefore, the reduced exposed human compartment is proximate to the sum of the two exposed human compartments of the full model when,

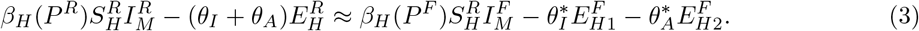

This condition implies that the latency rate of both non-immune and partially immune humans of the full model must be approximately the sum of diseased outflow rates from the exposed compartment of the reduced model. The solutions of the models are proximate when the conditions in Equations (2) and (3) both hold. To further explore the reduction of the full model to the reduced model, we will conduct a formal fitting analysis based on the parameters in the two conditions (see Section 3.2) and a parameter identifiability analysis on both models (see Section 3.1).

### 2.4 Parameter Identifiability Analysis

Parameter identifiability is an analytical tool that determines whether a model’s parameters can be uniquely estimated given some observed outputs [39, 28]. In this study, we explore two identifiability phenomena, structural (or a priori) and practical (or a posteriori) identifiability, as per Raue et al. (2009) and Gábor et al. (2017) [39, 40]. A model is said to be structurally identifiable if the parameters can be identified based on the model’s structure and equations, without considering specific data, whereas it is said to be practically identifiable if the parameters can be accurately estimated from data, using numerical methods and computational tools [39, 40]. The structural and practical identifiability analysis was conducted on both the full and reduced models to determine and compare the extent to which the parameters of each model can be identified from the model structure and noisy data.

By employing the StructuralIdentifiability.jl and SIAN.jl packages in Julia [41], we conducted the structural identifiability analysis under two scenarios. The first scenario considered all model compartments as outputs, and the second scenario considered a single output, namely the asymptomatic infectious human class, *A*_*H*_. We chose to observe the *A*_*H*_ class in the second scenario to reflect practical field studies where asymptomatic carriers are increasingly being detected through active case detection and serological surveys, making data for the asymptomatic infectious humans class readily available in real-world malaria surveillance [42, 43, 44, 45, 46, 47, 48, 49].

We further investigate the second scenario by simulating noisy data of the asymptomatic infectious human class *A*_*H*_ from each model and assessing practical identifiability. We simulated 180 data points while maintaining model parameters at baseline as indicated in Table 1, with a 5-day interval between consecutive data points and incorporating negative binomial noise. That is, for each of the 180 time points, *t*, we simulate data point, *A*_*Hnoise*_(*t*), from a negative binomial distribution with expected value *A*_*H*_ (*t*) and dispersion parameter *ϕ*, as presented by:

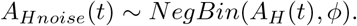

By employing Bayesian inference in STAN with 4000 iterations, we set Λ_*H*_, Λ_*M*_, *µ*_*H*_, *µ*_*M*_, *b, c, σ* and *σ*_2_ as per Table 1 and estimated the remaining 13 parameters of the reduced model and 14 parameters of the full model, investigating the effective sample size, standard error of the mean, and coherence amongst the 4 chains considered. The parameters Λ_*H*_, Λ_*M*_, *µ*_*H*_, *µ*_*M*_, *b, c, σ* and *σ*_2_ were fixed based on established range of demographic values, immunity waning rates and previously employed constant coefficients of the interventions class (*b* and *c*) allowing us to enhance the identifiability of the remaining parameters. Refer to Section 3.1 for the results of the structural and practical identifiability analyses.

## 3 Results

### 3.1 Parameter Identifiability

#### 3.1.1 Structural Identifiability

In the scenario where all model compartments are considered outputs, we observed that all parameters of both models are globally identifiable except for the mosquito biting rate (*δ*), and the transmission success parameter in humans (*ψ*_*H*_) and mosquitoes (*ψ*_*M*_), see Table 2. The identifiability issue with the parameters *δ, ψ*_*M*_ and *ψ*_*H*_ becomes obvious when the transmission rate functions are examined, as a model with parameters (*δ∗, ψ*_*M*_*∗*, *ψ*_*H*_*∗*) is indistinguishable from another model with (*δ*, *ψ*_*M*_, *ψ*_*H*_) = (1, *δ· ψ*_*M*_, *δ· ψ*_*H*_), yielding the same transmission rates. The result of this scenario indicates that both the full and reduced models exhibit the same structural identifiability when observing all model states as outputs. Similarly, in the scenario where only *A*_*H*_ is considered an output, we observed that all model states and parameters were identifiable, except for the mosquito biting rate (*δ*), the transmission success parameter in humans (*ψ*_*H*_) and mosquitoes (*ψ*_*M*_), constant coefficients of *P* in the human and mosquito transmission rate function (*b* and *c*), influx rate of interventions (*η*), relative infectiousness of asymptomatic humans (*ξ*), all mosquito states (*S*_*M*_, *E*_*M*_ and *I*_*M*_), and the *P* class (Table 2). The additional non-identifiable parameters and model states in this scenario result from the fact that these parameters and model states are not directly influenced by the observed model state, *A*_*H*_. Thus, both models exhibited the same structural identifiability characteristics under the two scenarios considered for the structural identifiability analysis.

**Table 2:** Summary of structural identifiability analysis conducted on the full and reduced models.

| Scenario | Identifiable parameters and states | Non-identifiable parameters and states |
| --- | --- | --- |
| All compartments as outputs | $\Lambda_H, \Lambda_M, \mu_H, \mu_M, \tau, \omega, \gamma, \nu, \chi,$<br>$\eta, \kappa, b, c, \xi, I_H, A_H, S_M, E_M, I_M, P$<br>Full model : $\theta_I^*, \theta_A^*, \sigma_1, \sigma_2,$<br>$S_{H1}, S_{H2}, E_{H1}, E_{H2}$<br>Reduced model : $\theta_I, \theta_A, \sigma, S_H, E_H$ | $\delta, \psi_H, \psi_M$ |
| Single output: $A_H$ | $\Lambda_H, \Lambda_M, \mu_H, \mu_M, \tau, \omega, \gamma, \nu, \chi,$<br>$\kappa, I_H, A_H$<br>Full model : $\theta_I^*, \theta_A^*, \sigma_1, \sigma_2,$<br>$S_{H1}, S_{H2}, E_{H1}, E_{H2}$<br>Reduced model : $\theta_I, \theta_A, \sigma, S_H, E_H$ | $\delta, \psi_H, \psi_M, b, c, \eta,$<br>$\xi, S_M, E_M, I_M, P$ |

#### 3.1.2 Practical Identifiability

Proceeding with the single output scenario, observing *A*_*H*_ for each model, we employed the baseline parameter values for Λ_*H*_, Λ_*M*_, *µ*_*H*_, *µ*_*M*_, *b, c, σ* and *σ*_2_ as per Table 1 while estimating the remaining parameters in each model, 13 parameters in the case of the reduced model and 14 for the full model. Noisey data were generated assuming negative binomial noise on simulated data from each model given the initial conditions *S*_*H*1_(0) = 3000, *S*_*H*2_(0) = 2000, *E*_*H*1_(0) = 150, *E*_*H*2_(0) = 50, *I*_*H*_ (0) = 5, *A*_*H*_ (0) = 10, *S*_*M*_ (0) = 1000, *E*_*M*_ (0) = 50, *I*_*M*_ (0) = 10, *P* (0) = 1 for the full model and *S*_*H*_ (0) = 5000, *E*_*H*_ (0) = 200, *I*_*H*_ (0) = 5, *A*_*H*_ (0) = 10, *S*_*M*_ (0) = 1000, *E*_*M*_ (0) = 50, *I*_*M*_ (0) = 10, *P* (0) = 1 for the reduced model, setting *θ*_*I*_ and *θ*_*A*_ to be same as 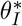 and 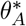 respectively for the reduced model simulation. Figure 3 compares the posterior distribution of parameters with the fixed values used to generate the data for each model, as stated in Section 2.4. The estimated parameters explored here are 13 parameters from the reduced model with the inverse dispersion parameter (*ϕ*^−1^), and likewise 14 parameters from the full model plus the inverse dispersion parameter. The results suggest that the parameters inferred here for both models are practically identifiable (refer to Figure 3 and Tables 6 and 7). This conclusion is drawn from several factors such as relatively small standard errors, large effective sample sizes, and the 95% credible intervals maintain appropriate widths, implying good precision with Rhat values close to 1 as found in Tables 6 and 7.

**Figure 3.**
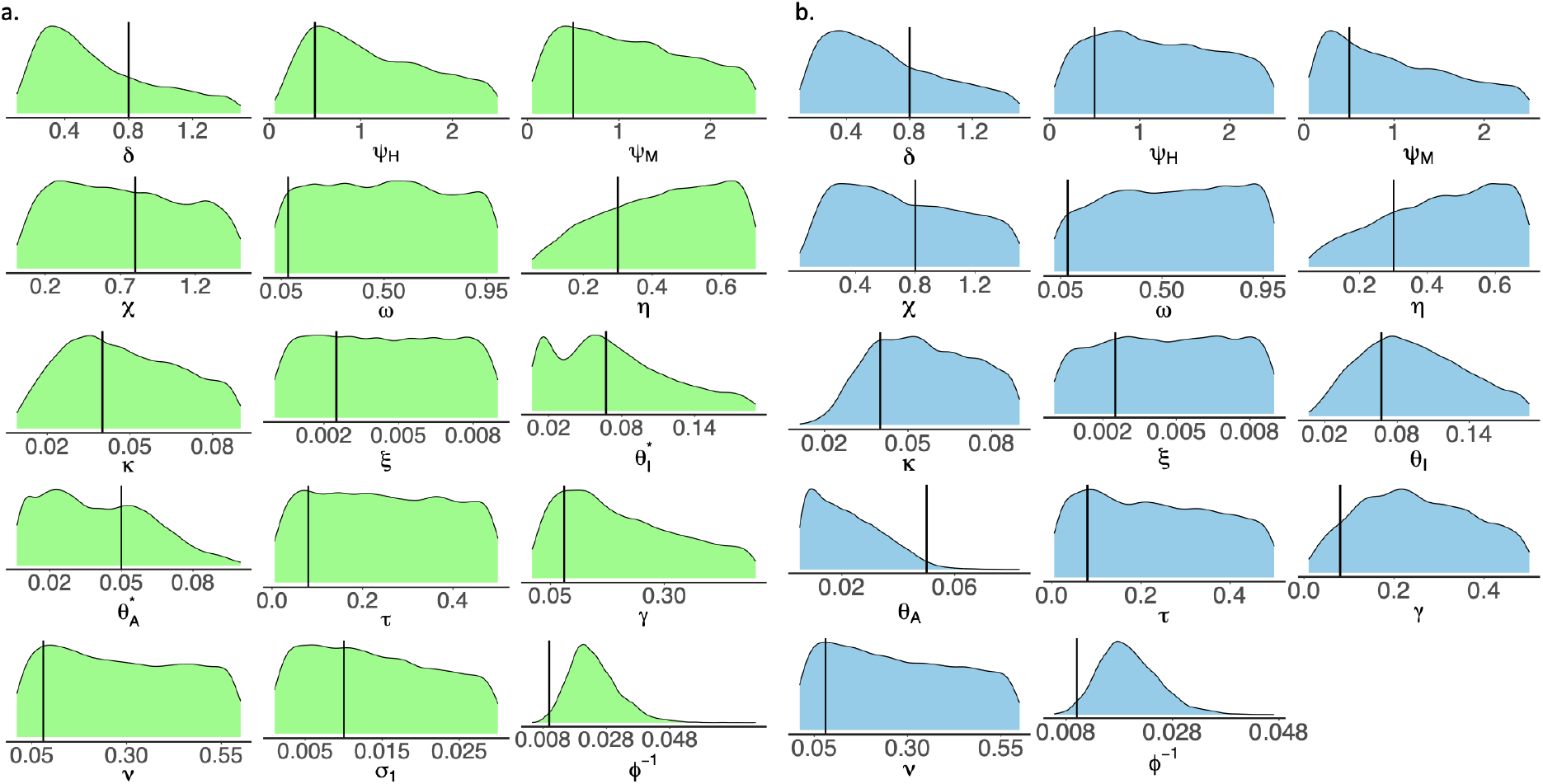
Marginal posterior density plots of parameters estimated in the practical identifiability analysis observing the single *A*_*H*_ output for both models. **a**. Results comparing the posterior densities of the full model parameters and the inverse dispersion parameter (*ϕ*^−1^) with the fixed values of the parameters utilised in the generation of data marked by the black lines. **b**. Results comparing the posterior densities of the reduced model parameters and the inverse dispersion parameter (*ϕ*^−1^) with the fixed values of the parameters utilised in the reduced model data simulation. The green-coloured plots correspond to results from the full model whereas the sky-blue-coloured plots correspond to results from the reduced model. The fixed value of the parameters utilised in the data simulation is marked by the black line.

We observe from Figure 3 and Tables 6 and 7 in Appendix Section B.3 that the marginal density functions have posterior support around the true parameter values used. Moreover, all of the true parameter values lie within the 95% credible intervals, see Figure 3, and Tables 6 and 7.

Comparing Tables 6 and 7, we observed that while there was broad agreement among the two sets of inferences generated, the parameters of the reduced model have comparatively higher effective sample sizes and lower standard mean errors compared to the full model. This indicates that the reduced model achieved faster convergence of estimates.

### 3.2 Bayesian MCMC Model-to-Model Fitting

To fit the reduced model to a more realistic simulation dataset of the full model, mimicking real-life epidemic scenarios with the incorporation of noise, Bayesian inference with Markov chain Monte Carlo (MCMC) was used. The MCMC algorithm used in this study explored the parameter space by iteratively generating samples to estimate the posterior distribution, enabling us to quantify the uncertainty associated with parameter estimates while deriving credible intervals. We conducted the full model to reduced model fitting analysis considering two scenarios; first with all of the full model compartments observed, see Section 3.2.1, and second with only the asymptomatic infectious compartment observed, refer to Section 3.2.2, as similarly considered in the practical identifiability analysis, refer to Section 2.4, since the second scenario reflects an increasingly practical data fitting scenario given that asymptomatic carriers are increasingly being detected in malaria surveillance studies.

#### 3.2.1 Fitting data from all full model compartments

Considering the baseline parameters in Table 1, we investigated the ability of the reduced model to provide solutions close to the full model by generating simulated data, *F* (*t*), with negative binomial noise from all 10 compartments of the full model. We then summed the two susceptible and the two exposed compartments of the full model to obtain a noisy dataset of eight compartments and fitted the reduced model to the resulting data. We suppose that the 10 compartments of the full model are observed at 5-day intervals over a 4-year period. The observed data, *F* (*t*), were generated by assuming negative binomial noise on the simulated data from the full model given the initial conditions *S*_*H*1_(0) = 3000, *S*_*H*2_(0) = 2000, *E*_*H*1_(0) = 150, *E*_*H*2_(0) = 50, *I*_*H*_ (0) = 5, *A*_*H*_ (0) = 10, *S*_*M*_ (0) = 1000, *E*_*M*_ (0) = 50, *I*_*M*_ (0) = 10, *P* (0) = 1 and the parameter values from Table 1. These parameters give 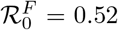. The observed data from the full model, *F* (*t*), have a negative binomial likelihood. Similarly, we consider data from the reduced model, *R*(*t*), to have a negatively binomially distributed likelihood with the expected value *R*(*t*) and dispersion parameter *ϕ*: *F* (*t*) *~* NegBin(*R*(*t*), *ϕ*).

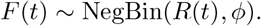

We compare the dynamics of the models by fitting the reduced model to the observed data from the full model, estimating posterior distribution for the model parameters, *θ* = (*θ*_*I*_, *θ*_*A*_, *σ, τ, ϕ*^−1^), and making posterior predictive checks. We define the following prior distributions for the five parameters

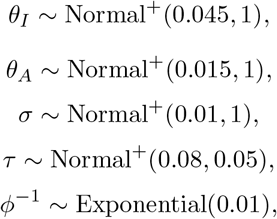

employing parameter estimates from a least squares fit as the mean of the priors. Normal^+^ refers to the normal distribution truncated at zero, where all negative values are excluded. We define the dispersion parameter, *ϕ*, using *ϕ*^−1^ as this reparameterisation helps prevent the prior from concentrating too much mass on models exhibiting high overdispersion [50, 51].

In Figure 4, we observe from the posterior predictive check that the reduced model fits the simulated data of the full model well, as the smooth simulations and noisy data are both captured within the 95% credible intervals. We observe from Table 3 that Rhat is 1, suggesting a high level of agreement among the four Markov chains, which is further confirmed in Figure 9. Additionally, the effective sample size (“n eff”) is reasonably high, indicating that the Markov chains effectively explored the parameter space cohesively [50]. In comparing the prior and posterior distributions in Figure 9, we observe substantial parameter learning across all model parameters. Posteriors for *θ*_*I*_ and *θ*_*A*_ exhibit moderate concentration with slight shifts from prior means, while *σ, τ*, and particularly *ϕ*^−1^ demonstrate pronounced posterior concentration. In Figure 5, we observe a positive correlation between *σ* and *θ*_*A*_, negative correlations between *θ*_*I*_ and *θ*_*A*_, *σ* and *τ*, and *θ*_*I*_ and *σ*. These posterior behaviours are not surprising given the combined nature of the immunity pathways in the reduced model (considered separate in the full model), see Figure 2. For instance, the negative correlation between *σ* and *τ* indicates that as symptomatic individuals receive more treatment (rate parameter, *τ*), the immunity waning removal from the *A*_*H*_ class must decline (rate parameter, *σ*) for fixed observed data, see Figure 2. Using the same approach, we also explore the proximity of the reduced model to the full model for a high malaria transmission scenario with 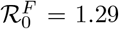, see Appendix C.1. We now proceed with the model-to-model fitting analysis assuming a more realistic, limited observed data, scenario for the low malaria transmission scenario in Section 3.2.2.

**Table 3:** Summary of Bayesian parameter inference obtained from the model-to-model fitting considering a combined dataset from all compartments of the full model and assuming a low transmission scenario with 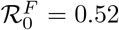. Posterior estimates shown for the reduced model parameters *θ*_*I*_, *θ*_*A*_, *σ* and *τ*, as well as the inverse dispersion parameter *ϕ*^−1^.

| Parameters | mean | se_mean | sd | 2.5% | 25% | 50% | 75% | 97.5% | n_eff | Rhat |
| --- | --- | --- | --- | --- | --- | --- | --- | --- | --- | --- |
| $\theta_I$ | 0.041 | 0.00003 | 0.0018 | 0.037 | 0.039 | 0.041 | 0.042 | 0.044 | 4055 | 1 |
| $\theta_A$ | 0.018 | 0.00002 | 0.0015 | 0.0015 | 0.017 | 0.018 | 0.019 | 0.021 | 4055 | 1 |
| $\sigma$ | 0.012 | 0.000005 | 0.0003 | 0.0115 | 0.0119 | 0.0120 | 0.0122 | 0.013 | 3235 | 1 |
| $\tau$ | 0.042 | 0.00014 | 0.009 | 0.0259 | 0.036 | 0.042 | 0.048 | 0.061 | 4326 | 1 |
| $\phi^{-1}$ | 0.011 | 0.00001 | 0.0007 | 0.0095 | 0.0103 | 0.0107 | 0.0112 | 0.012 | 6232 | 1 |

**Figure 4.**
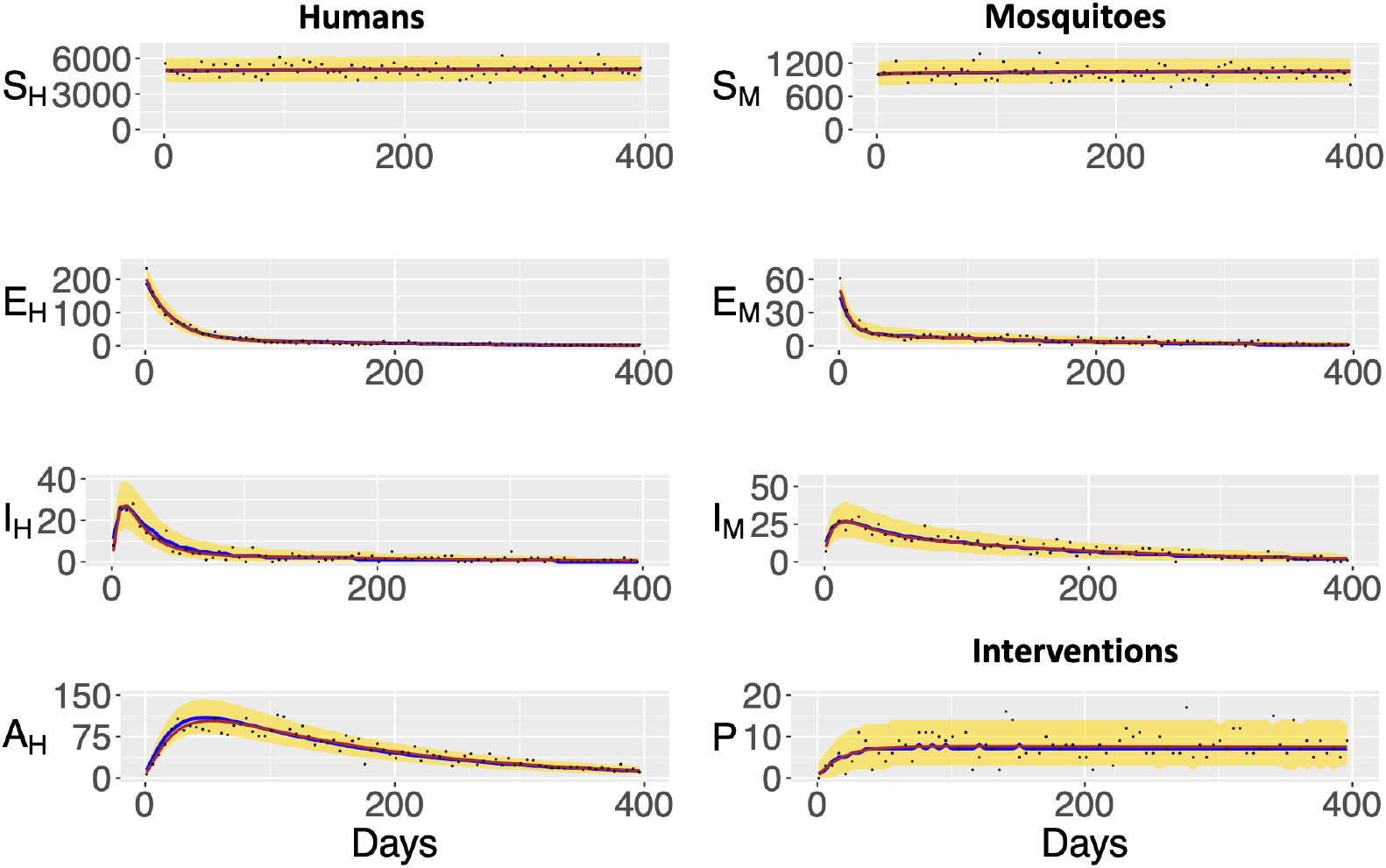
Posterior predictive check of the compartments in the reduced model. This figure visualises the model fitting dynamics up to 400 days, as the model stabilises to the disease-free equilibrium within this timeframe, and extending to the full timeframe, 1400 days would make the plots difficult to interpret. The blue solid line is the median and the yellow area is the associated 95% credible intervals. The black points represent the noisy simulated data generated from the full model and the brown solid lines are the (combined) full model simulation without noise. These results were obtained assuming a low malaria transmission scenario with the full model at 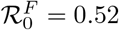 and the reduced model at 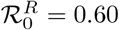 (using the mean estimates of *θ*_*I*_, *θ*_*A*_, *σ* and *τ* from Table 3).

**Figure 5.**
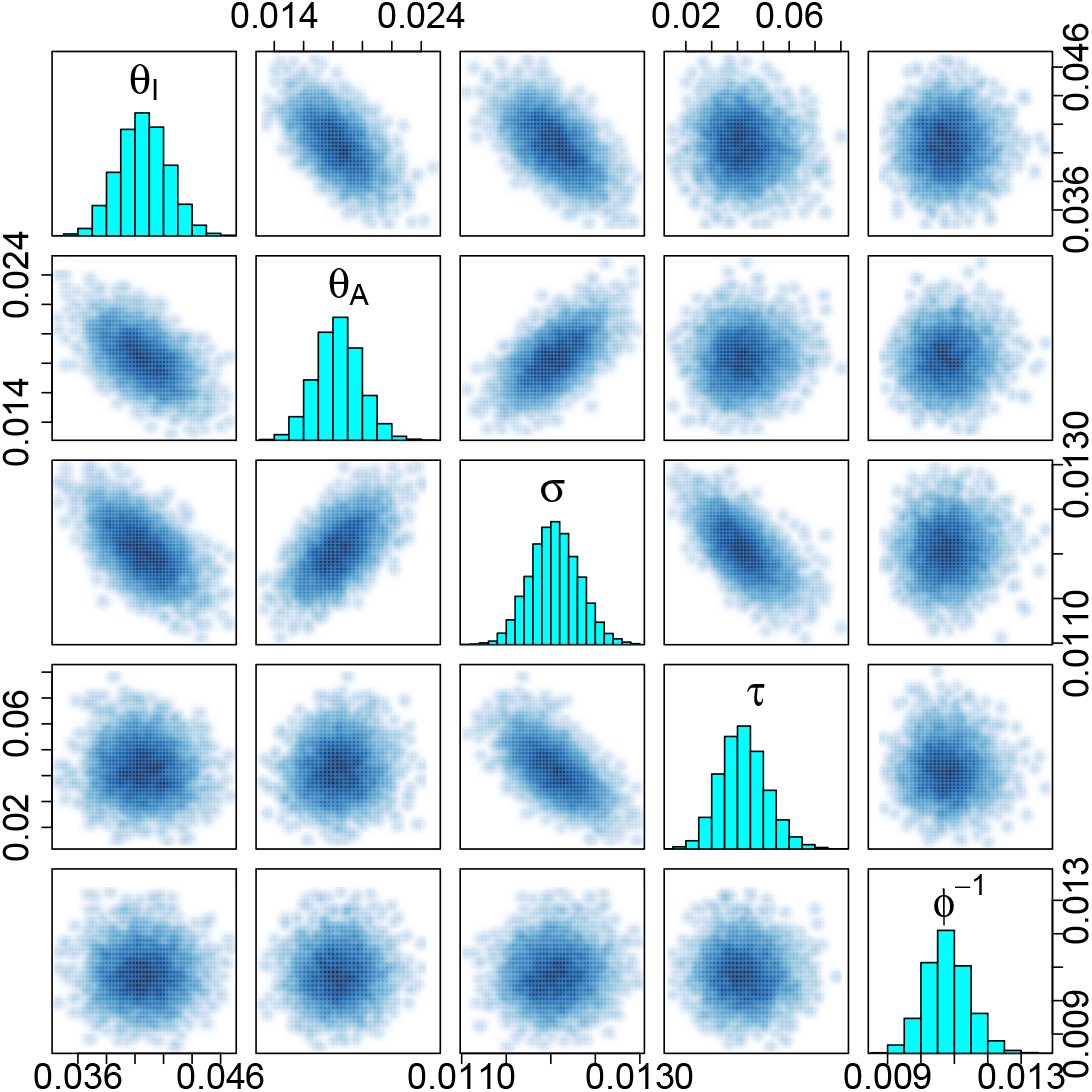
Pairwise plots of estimated reduced model parameters *θ*_*I*_, *θ*_*A*_, *σ* and *τ*, and the inverse dispersion parameter *ϕ*^−1^ for the model-to-model inference, assuming a low transmission scenario with 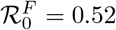 and 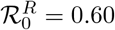 (using the mean estimates of *θ*_*I*_, *θ*_*A*_, *σ* and *τ* from Table 3).

#### 3.2.2 Fitting data from asymptomatic infectious humans *A*_*H*_ only

In this scenario, we assume that the only observable data is the number of asymptomatic infections in the population, measured every five days across a four-year period. We consider the prevalence of asymptomatic infections in this study, as they are increasingly the focus of research due to their contribution to malaria transmission in endemic areas [42, 43, 44, 45, 46, 47, 48, 49]. Given the low transmission scenario, and the noise-incorporated simulation data generated from the full model in Section 3.2.1, we only employ data from the asymptomatic infectious class of the full model. We observed the full model through an observational model where the asymptomatic infectious class of the reduced model, *A*_*H*_ (*t*) is linked to the observed data of the full model’s asymptomatic infectious class *A*_*OBS*_(*t*) at each time point. We represent *A*_*OBS*_(*t*) with a negative binomial distribution, enabling us to utilise *A*_*H*_ (*t*) as the expected value while accommodating over-dispersion by incorporating the parameter *ϕ*, as presented below;

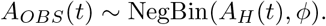

We fit the reduced model’s asymptomatic class to the observed data to obtain a posterior distribution for the model parameters, *θ* = (*θ*_*I*_, *θ*_*A*_, *σ, τ, ϕ*^−1^). The prior distributions for each of the five parameters are the same as defined in Section 3.2.1.

In Figure 6, we observe from the posterior predictive check that the asymptomatic infectious class of the reduced model fits the actual simulated data of the full model (brown solid line) as well as the noise-incorporated data (black points) fairly well as the noisy data and smooth simulated data are both captured within the 95% interval in the yellow shade. We also observe a high level of agreement among the four Markov chains from Table 4, which is further confirmed by observing the marginal posterior density plots in Figure 10. In Figure 10, we observe that the posterior densities compared to the previous distributions are more concentrated across all parameters, particularly for *θ*_*A*_, and *σ*, reflecting increased precision, strongly informed by data. In comparing the inference tables, Tables 3 and 4, we observe that while there is agreement amongst the four chains considered in two fitting scenarios, the estimated parameters of the reduced model have comparatively higher effective sample sizes and lower standard mean errors in the scenario where all compartments of the full model are observed than in the scenario with limited data and only the asymptomatic class, *A*_*H*_ of the full model is observed. This indicates that more reliable parameter estimations as well as a more accurate fit were obtained in the scenario where all compartments of the full model are observed. Moreover, the complex (observed between *θ*_*A*_ and *σ, θ*_*A*_ and *θ*_*I*_, and *σ* and *τ*), non-linear (observed between *θ*_*I*_ and *σ*) correlations and the identifiability issues observed in the correlation plots in Figure 7 as compared to Figure 5 can be attributed to the limited data (only the asymptomatic class, *A*_*H*_ of the full model) observed. We explore a similar analysis for a high malaria transmission situation with 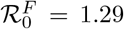 employing a similar approach in Appendix C.1.

**Table 4:** Summary of Bayesian parameter inference table obtained from the model-to-model fitting considering data from only the asymptomatic class of the full model, assuming a low transmission situation with 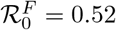. The table provides posterior estimates for the reduced model’s parameters *θ*_*I*_, *θ*_*A*_, *σ* and *τ*, as well as the inverse dispersion parameter *ϕ*^−1^.

| Parameters | mean | se.mean | sd | 2.5% | 25% | 50% | 75% | 97.5% | n_eff | Rhat |
| --- | --- | --- | --- | --- | --- | --- | --- | --- | --- | --- |
| $\theta_I$ | 0.011 | 0.00015 | 0.00696 | 0.00059 | 0.00621 | 0.0109 | 0.0159 | 0.0266 | 2015 | 1 |
| $\theta_A$ | 0.016 | 0.00004 | 0.0018 | 0.0128 | 0.0153 | 0.0165 | 0.0176 | 0.02 | 2359 | 1 |
| $\sigma$ | 0.015 | 0.00005 | 0.00197 | 0.0125 | 0.0138 | 0.0149 | 0.0166 | 0.0197 | 1535 | 1 |
| $\tau$ | 0.084 | 0.00081 | 0.0439 | 0.0096 | 0.0502 | 0.0815 | 0.113 | 0.177 | 2910 | 1 |
| $\phi^{-1}$ | 0.026 | 0.00013 | 0.0082 | 0.0126 | 0.0202 | 0.0251 | 0.0310 | 0.0444 | 3755 | 1 |

**Figure 6.**
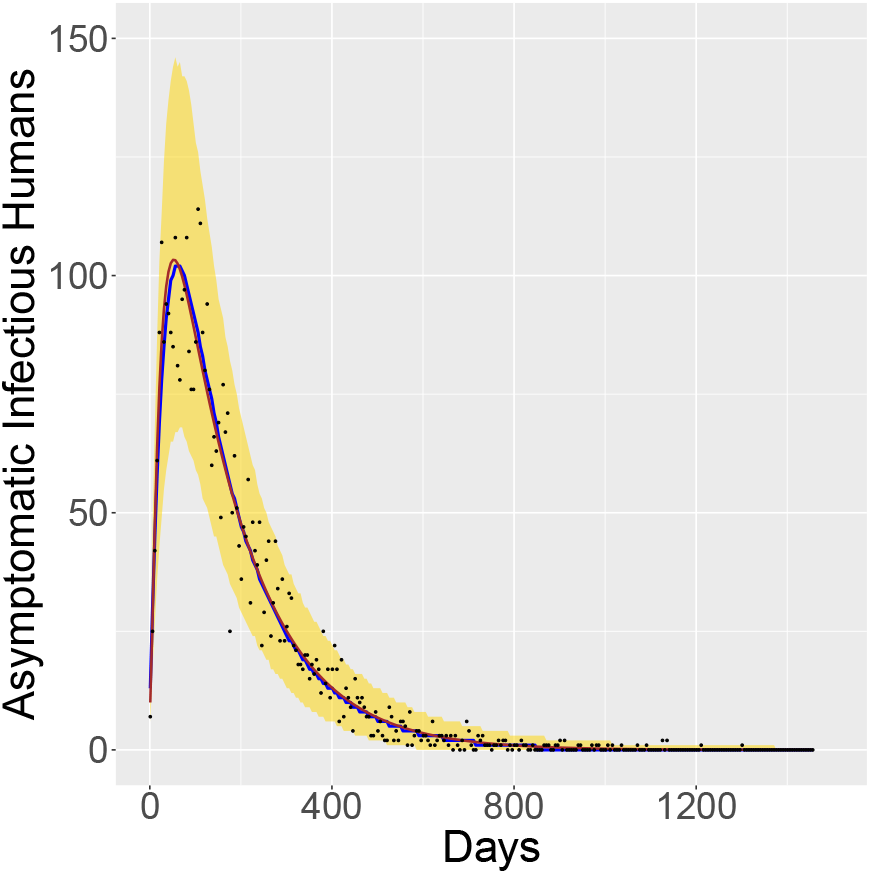
Posterior predictive check of the asymptomatic infectious compartment of the reduced model. The blue solid line is the median line and the yellow region, the 95% credible interval of the reduced model’s asymptomatic infectious compartment. The black points represent the noise-incorporated data points generated from the full model whereas the brown solid line represents the smooth simulation data generated from the full model without noise. This result was obtained assuming a low malaria transmission situation with the full model at 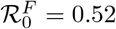 and the reduced model at 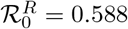 (using the mean estimates from Table 4).

**Figure 7.**
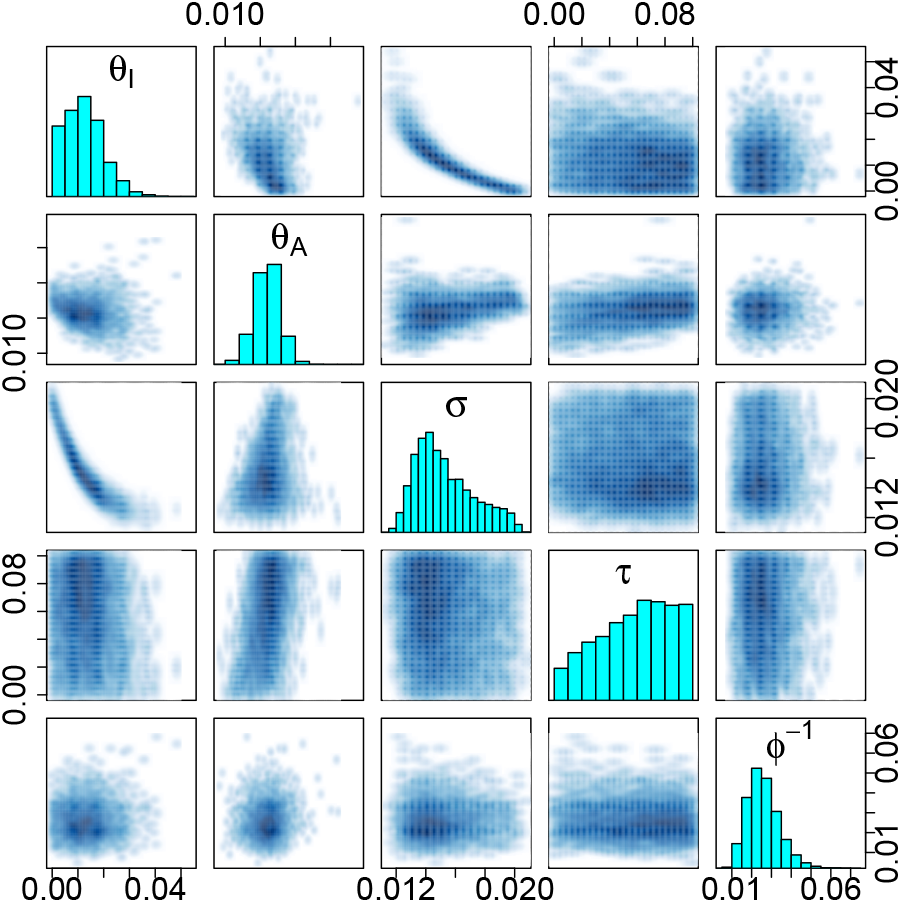
Pairwise plots of the estimated reduced model parameters; *θ*_*I*_, *θ*_*A*_, *σ* and *τ*, and the inverse dispersion parameter *ϕ*^−1^ from the inference generated from the model-to-model fitting analysis with observing only the *A*_*H*_ class, assuming a low transmission scenario with 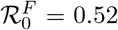 and 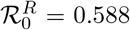 (using the mean estimates from Table 4).

Based on the results presented in Sections 3.2.1 and 3.2.2, we conclude that the reduced model serves as a good approximation of Korsah et al.’s full model, demonstrating similar transmission characteristics as the full model while being more adaptable for further studies across several endemic settings. Importantly, our results from Section 3.2 show that we can make meaningful predictions about the reduced model when fitting to a realistic dataset (noise-incorporated and limited data) generated from the full model.

## 4 Discussion

In this paper, we simplify the comprehensive malaria control model by Korsah et al. into a reduced system that retains similar transmission characteristics of the original model by combining the separated partially immune and non-immune human pathways of the original model [26]. We do this to address the practical challenge of observing partially immune and non-immune individuals in a population, especially in endemic regions where serological markers of immunity show wide variation and are not always available [30, 31]. We analysed the reduction technique employed in simplifying the full model into the reduced model (Section 2.3), conducted parameter identifiability analysis (Section 2.4) comparing both systems and conducted model-to-model fitting analyses employing Bayesian MCMC fitting approach (Sections 3.2 and C.1). Our findings demonstrate that, although the reduced model does not explicitly model the separate transmission routes of the two immunity levels in the human population as captured in the full model, it is sufficient for decision making and future model extensions. Moreover, the reduced system provides a simplified and proximate framework to inform decisions, potentially helping to improve progress toward WHO 2030 goals, as outlined in the Global Technical Strategy [7, 8].

The assessment of the basic reproduction number of the reduced model identified three pathways of transmission; *R*_1_ represents transmissions from symptomatic infectious individuals in *I*_*H*_ (Equation (9)), *R*_2_ represents transmissions from individuals with asymptomatic second infections (Equation (10)) in *A*_*H*_, and *R*_3_ considers transmissions from self-recovered individuals (symptomatic infectious individuals) in the *A*_*H*_ class (Equation (11)), see Appendix B.1. The inclusion of the *R*_2_ transmission route in the reduced model (but not the full model; see Appendix A) gives policymakers the opportunity to utilize the *R*_2_ formulation estimates to control malaria transmissions from asymptomatic individuals with asymptomatic second infections. Effectively targeting this transmission route requires stakeholders to first quantify its impact by estimating *R*_2_ based on population-specific dynamics. With a high *R*_2_ estimate, public health officials can deploy targeted interventions including Mass Screening and Treatment (MSaT), Mass Drug Administration (MDA), or Focused Screening and Treatment (FSaT) to specifically disrupt transmission originating from these asymptomatic carriers [52, 53]. This model-guided approach enables a more efficient allocation of limited public health resources to interventions that specifically address this often overlooked transmission pathway.

A key strength of the work presented in this study is the rigorous validation of the reduced model through the analysis of the reduction technique employed, parameter identifiability, sensitivity and model-to-model fitting analyses using Bayesian MCMC fitting approach, all of which demonstrated the closeness of the models, see Section 3. We particularly obtained the same results in the structural identifiability analysis of both models and similar dynamics were observed for the practical identifiability analysis. However, we derived more accurate and reliable estimates for the reduced model’s parameters than the full model in our practical identifiability results, see Section 3.1. The similarity observed for both models likely results from the reduction technique employed, which preserved the essential transmission dynamics while decreasing the full model’s complexity from 10 states and 21 parameters to 8 states and 20 parameters, see Figures 1 and 2. The sensitivity analysis conducted on the reduced model confirms previous findings that parameters such as the mosquito biting rate (*δ*) and infection success rates (*ψ*_*H*_, *ψ*_*M*_) positively influence ℛ_0_ whereas parameters like the treatment rate of infected persons (*τ*) and mosquito death rate (*µ*_*M*_) negatively impact ℛ_0_, refer to Appendix B.2 [26, 16, 32]. Importantly, we identified parameters such as the clinical infection recovery rate (*γ*) and the probability of clinical infections in non-immune humans after latency (*ρ*) whose sensitivity was dependent on the relative infectiousness of asymptomatic humans (*χ*), see Appendix B.2 and Table 5. These sensitivity relations can be strategically adjusted to help control malaria outbreaks, offering policymakers specific targets for intervention programs in endemic regions. For instance, policymakers of a population with a high *χ* (relative infectiousness of asymptomatic humans) value can control malaria outbreaks by promoting malaria treatment campaigns, decreasing the self-recovery rate parameter (*γ*).

**Table 5:** Sensitivity index of the reduced model parameters on ℛ_0_ and 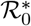.

| Parameters | Sensitivity index on $\mathcal{R}_0$ and $\mathcal{R}_0^*$ |
| --- | --- |
| $\delta$ | 1 |
| $\psi_H$ | 0.5 |
| $\psi_M$ | 0.5 |
| $\omega$ | $\frac{\mu_M}{2(\mu_M + \omega)} > 0$ |
| $\chi$ | $\frac{1}{2[1 + \frac{\rho(\mu_H + \sigma)}{\chi((1-\rho)(\tau + \nu + \mu_H) + \gamma)}]} > 0$ |
| $\theta$ | $\frac{\mu_H}{2(\mu_H + \theta)} > 0$ |
| $\rho$ | $\frac{1}{2[1 + \frac{\chi(\tau + \nu + \mu_H + \gamma)}{\rho((\mu_H + \sigma) - \chi(\tau + \nu + \mu_H))}]} > 0$, if $\frac{\mu_H + \sigma}{\tau + \nu + \mu_H} > \chi$ |
| $\gamma$ | $\frac{\gamma[1 - \chi(\tau + \nu + \mu_H)]}{2(\tau + \nu + \mu_H + \gamma)[(\mu_H + \sigma) + \chi(\frac{(1-\rho)}{\rho} + \gamma)]} < 0$, if $\chi > \frac{1}{(\tau + \nu + \mu_H)}$ |
| $\mu_M$ | $-\frac{3\mu_M + 2\omega}{2(\mu_M + \omega)} < 0$ |
| $\tau$ | $-\frac{\tau}{2(\tau + \nu + \mu_H + \gamma)[1 + \frac{(\tau + \nu + \mu_H + \gamma)\chi(1-\rho)}{\rho(\mu_H + \sigma + \chi\gamma)}]} < 0$ |
| $\nu$ | $-\frac{\nu}{2(\tau + \nu + \mu_H + \gamma)[1 + \frac{(\tau + \nu + \mu_H + \gamma)\chi(1-\rho)}{\rho(\mu_H + \sigma + \chi\gamma)}]} < 0$ |
| $\sigma$ | $-\frac{\sigma}{2(\mu_H + \sigma)[1 + \frac{(\mu_H + \sigma)\rho}{\chi((1-\rho)(\tau + \nu + \mu_H) + \gamma)}]} < 0$ |
| Intervention Parameters | Sensitivity index on $\mathcal{R}_0$ |
| $\kappa$ | $\frac{\eta[2bc\eta + (b+c)\kappa]}{2(b\eta + \kappa)(c\eta + \kappa)} > 0$ |
| $b$ | $-0.5(1 + b\eta) < 0$ |
| $c$ | $-0.5(1 + c\eta) < 0$ |
| $\eta$ | $-\frac{\eta[2bc\eta + (b+c)\kappa]}{2(b\eta + \kappa)(c\eta + \kappa)} < 0$ |

**Table 6:** Bayesian inference for reduced model generated from STAN. Four chains were used, each with 4000 iterations (burn-in of 2000). For each parameter, we present the posterior mean, the standard error of the mean (se mean), the posterior standard deviation (sd), the 2.5%, 25%, 50% (median), 75%, and 97.5% percentiles of the posterior distribution, (with the 2.5% and 97.5% percentiles forming the bounds of the 95% credible interval) the effective sample size (n eff) and the convergence diagnostic (Rhat).

| Parameters | mean | se_mean | sd | 2.5% | 25% | 50% | 75% | 97.5% | n_eff | Rhat |
| --- | --- | --- | --- | --- | --- | --- | --- | --- | --- | --- |
| $\delta$ | 0.646 | 0.004 | 0.359 | 0.146 | 0.353 | 0.577 | 0.901 | 1.409 | 6636 | 1 |
| $\psi_H$ | 1.220 | 0.009 | 0.678 | 0.137 | 0.647 | 1.173 | 1.790 | 2.421 | 6104 | 1 |
| $\psi_M$ | 1.030 | 0.008 | 0.681 | 0.106 | 0.431 | 0.911 | 1.556 | 2.388 | 7673 | 1 |
| $\chi$ | 0.718 | 0.004 | 0.398 | 0.112 | 0.377 | 0.671 | 1.046 | 1.448 | 7882 | 1 |
| $\omega$ | 0.539 | 0.003 | 0.280 | 0.048 | 0.304 | 0.546 | 0.782 | 0.980 | 9402 | 1 |
| $\eta$ | 0.442 | 0.002 | 0.169 | 0.100 | 0.314 | 0.460 | 0.586 | 0.689 | 8517 | 1 |
| $\kappa$ | 0.056 | 0.0002 | 0.018 | 0.025 | 0.041 | 0.055 | 0.070 | 0.088 | 8029 | 1 |
| $\xi$ | 0.005 | 0.00003 | 0.003 | 0.0003 | 0.002 | 0.005 | 0.007 | 0.009 | 10565 | 1 |
| $\theta_I$ | 0.095 | 0.0006 | 0.043 | 0.023 | 0.061 | 0.091 | 0.126 | 0.180 | 4850 | 1 |
| $\theta_A$ | 0.022 | 0.00015 | 0.012 | 0.006 | 0.012 | 0.020 | 0.030 | 0.048 | 7026 | 1 |
| $\tau$ | 0.235 | 0.001 | 0.142 | 0.016 | 0.109 | 0.226 | 0.354 | 0.486 | 10442 | 1 |
| $\gamma$ | 0.253 | 0.002 | 0.124 | 0.039 | 0.154 | 0.244 | 0.351 | 0.483 | 6847 | 1 |
| $\nu$ | 0.279 | 0.002 | 0.170 | 0.020 | 0.130 | 0.264 | 0.423 | 0.579 | 8504 | 1 |
| $\phi^{-1}$ | 0.020 | 0.00005 | 0.006 | 0.010 | 0.016 | 0.019 | 0.023 | 0.033 | 11094 | 1 |

**Table 7:** Bayesian inference for full model generated from STAN. Four chains were used, each with 4000 iterations (burn-in of 2000). The columns presented here are same as defined in Table 6.

| Parameters | mean | se_mean | sd | 2.5% | 25% | 50% | 75% | 97.5% | n_eff | Rhat |
| --- | --- | --- | --- | --- | --- | --- | --- | --- | --- | --- |
| $\delta$ | 0.610 | 0.006 | 0.352 | 0.151 | 0.329 | 0.515 | 0.834 | 1.405 | 3032 | 1 |
| $\psi_H$ | 1.132 | 0.011 | 0.644 | 0.193 | 0.587 | 1.023 | 1.632 | 2.404 | 3407 | 1 |
| $\psi_M$ | 1.150 | 0.010 | 0.679 | 0.120 | 0.563 | 1.086 | 1.699 | 2.412 | 4216 | 1 |
| $\chi$ | 0.729 | 0.006 | 0.411 | 0.076 | 0.376 | 0.705 | 1.074 | 1.454 | 4438 | 1 |
| $\omega$ | 0.504 | 0.004 | 0.282 | 0.042 | 0.261 | 0.503 | 0.741 | 0.977 | 5150 | 1 |
| $\eta$ | 0.435 | 0.003 | 0.169 | 0.104 | 0.304 | 0.452 | 0.579 | 0.687 | 4342 | 1 |
| $\kappa$ | 0.048 | 0.0003 | 0.021 | 0.013 | 0.031 | 0.046 | 0.065 | 0.087 | 3577 | 1 |
| $\xi$ | 0.004 | 0.00003 | 0.003 | 0.0003 | 0.002 | 0.004 | 0.007 | 0.009 | 5890 | 1 |
| $\theta_I^*$ | 0.074 | 0.001 | 0.046 | 0.010 | 0.037 | 0.067 | 0.104 | 0.175 | 2288 | 1 |
| $\theta_A^*$ | 0.040 | 0.001 | 0.022 | 0.008 | 0.021 | 0.038 | 0.057 | 0.087 | 1505 | 1 |
| $\sigma_1$ | 0.014 | 0.0001 | 0.008 | 0.002 | 0.007 | 0.014 | 0.021 | 0.029 | 4297 | 1 |
| $\tau$ | 0.249 | 0.002 | 0.143 | 0.019 | 0.125 | 0.245 | 0.372 | 0.488 | 3906 | 1 |
| $\gamma$ | 0.211 | 0.003 | 0.132 | 0.025 | 0.100 | 0.186 | 0.310 | 0.479 | 2469 | 1 |
| $\nu$ | 0.291 | 0.003 | 0.172 | 0.023 | 0.138 | 0.281 | 0.440 | 0.583 | 4593 | 1 |
| $\phi^{-1}$ | 0.024 | 0.0001 | 0.008 | 0.011 | 0.019 | 0.023 | 0.029 | 0.042 | 5709 | 1 |

Despite the promising results of obtaining a simplified version of Korsah et al.’s model, several questions remain unanswered regarding the extension of the reduced model to real-life data and the impact of utilising multiple interventions. In the future, an expansion of this work is recommended to fit the reduced model to data from a malaria-endemic setting. Future studies can also expand the reduced model to explore the impact of applying multiple concurrent interventions in an endemic area. Although robust, the model-to-model fitting conducted in this study utilises only simulated data and does not include validation against field data. We therefore recommend expanding this work to fit both models to data from malaria endemic settings to obtain parameter estimates to accurately parameterise the models and to establish a meaningful comparison between the two models against field data. Establishing this will better inform the use of the reduced model as to when it deviates from the full model.

In conclusion, this study demonstrates that reducing the complexity of a model while retaining key features can enhance its effectiveness in fitting diverse datasets and improve the accuracy of parameter estimations, since the reduced model explored in this study successfully maintains the essential dynamics of the full model while offering simplicity and improved parameter estimation. Our analysis demonstrates that the reduced model is sufficient for decision-making and can serve as a foundation for further model development, effectively replacing the full model. Particularly, our findings in Section 3.2 demonstrate that the reduced model can produce meaningful predictions about the full model’s transmission dynamics when fitted to noise-incorporated and limited datasets derived from the full model. Therefore, we recommend the use of the reduced system for future studies, as the reduced model allows stakeholders to better manage asymptomatic second infections in a population through the *R*_2_ transmission route, and can provide more reliable parameter estimates when fitted to data. As global efforts continue to meet the WHO 2030 malaria targets, this reduced framework offers a practical yet simplified approach to inform and optimise malaria control strategies.

## 5 Author Contributions

M. A. Korsah, S. T. Johnston, C. R. Walker and J. A. Flegg conceived the study. M. A. Korsah conducted the simulations and drafted the manuscript. S. T. Johnston, C. R. Walker, K. E. Tiedje and K. P. Day critically reviewed and revised the manuscript for intellectual content. J. A. Flegg oversaw project coordination and contributed to the manuscript revisions. All authors read and approved the final manuscript.

## 6 Acknowledgments

We acknowledge the support of the Melbourne Research Scholarship awarded to M. A. Korsah.

## 8 Data Availability

The data supporting the findings of this study are openly available on GitHub at https://github.com/AkuaK/Amalaria-control-model-reduction-and-analysis. We are currently refining the code and will ensure it is fully documented.

## A A Full Malaria Control Model

The governing equations for the Korsah et al. (2024) malaria control model are:

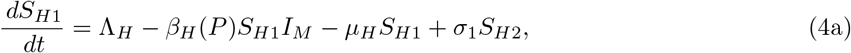

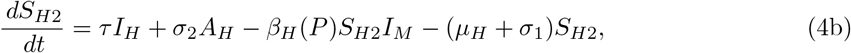

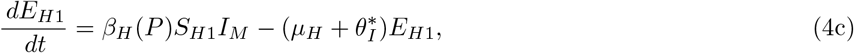

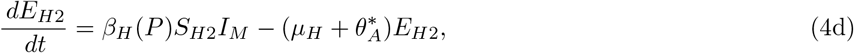

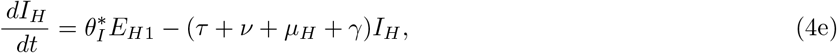

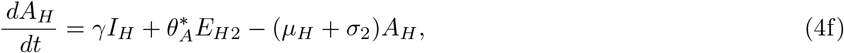

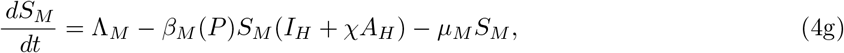

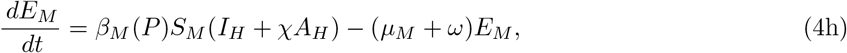

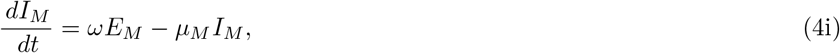

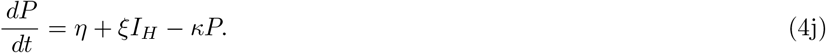

The basic reproduction number of the full model [26] was formulated as,

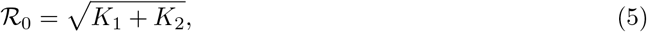

where *K*_1_ and *K*_2_ are defined as;

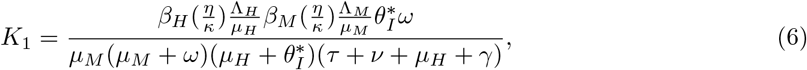

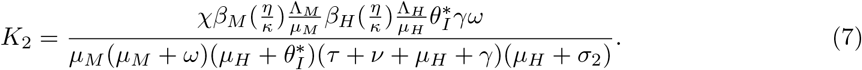

From ℛ_0_ in Equation (5), we identify two transmission links;

- *K*_1_ represents transmission from individuals in *I*_*H*_,
- *K*_2_ accounts for transmission occurring from individuals in *A*_*H*_ who have recovered from the *I*_*H*_ class without medical intervention, yet remain infectious despite showing no clinical symptoms.

Transmission from infected mosquitoes, *I*_*M*_, is accounted for in each transmission pathway, *K*_1_ and *K*_2_ [26].

## B Reduced Model Analysis

### B.1 Estimation of the basic reproduction number of the reduced model

The basic reproduction number is formulated using the next generation method [54] by first finding the new infection matrix:

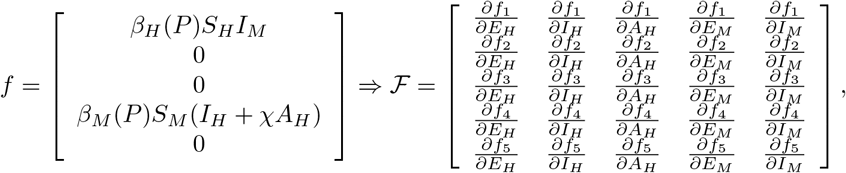

which gives

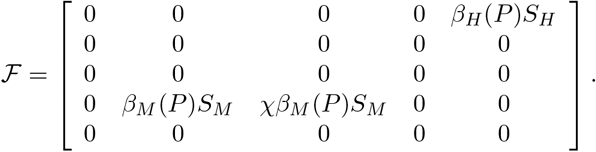

And the transition matrix

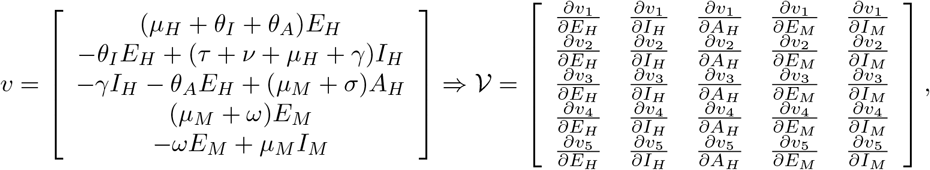

which gives

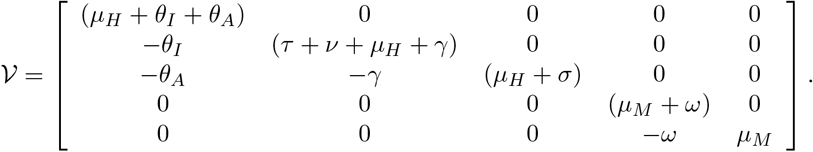

We compute ℛ_0_ by finding the spectral radius, *ρ*, of ℱ𝒱^−1^

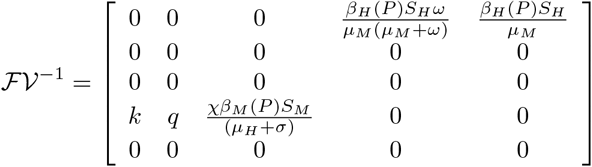

from the five infected compartments [*E*_*H*_, *I*_*H*_, *A*_*H*_, *E*_*M*_, *I*_*M*_] as detailed in the model equations, where

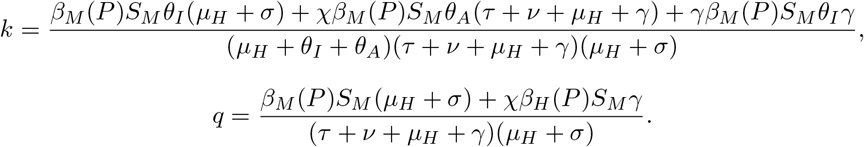

We consider the model at the diseases free equilibrium (DFE) 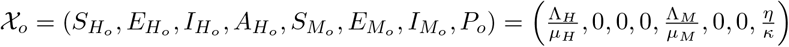. Thus,

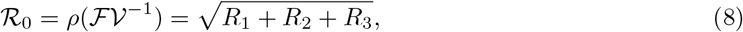

where *R*_1_, *R*_2_, and *R*_3_ are

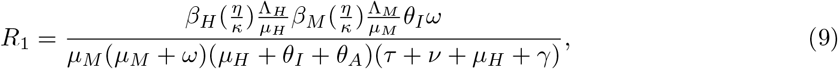

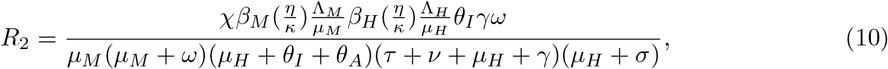

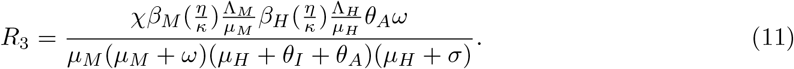

We identify three transmission pathways from the human population observed from ℛ_0_ in Equation (B.1);

- *R*_1_ represents transmissions from clinically infectious individuals in *I*_*H*_
- *R*_2_ represents transmissions from self-recovered infectious individuals in *A*_*H*_
- *R*_3_ represents transmissions from partially immune asymptomatic individuals, from *E*_*H*_ to *A*_*H*_

In comparing the transmission routes explored by the reproduction numbers of both models (see Appendix A), it is noticed that the *R*_1_ and *R*_3_ pathways of the reduced model are similarly represented in the full model as *K*_1_ and *K*_2_, respectively, refer to Appendix A and B.1 [26]. However, the *R*_2_ pathway is not considered in the full model as the full model in its complexity does not consider transmission from second infections at the DFE, an equilibrium state considered for the formulation of ℛ_0_ using the next generation method. Notice that in the absence of intervention programs (*P* = 0) the basic reproduction number becomes:

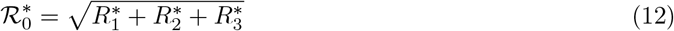

Where 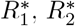, and 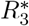 are

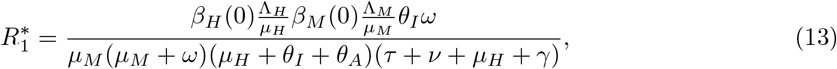

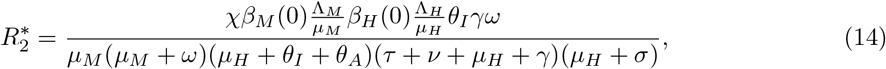

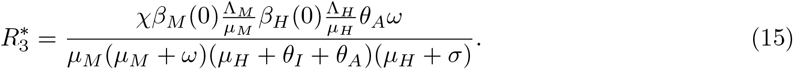

In referring to the model assumptions of Korsah et al. (2024), we observe that

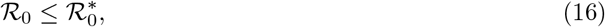

which is a similar result obtained for the full model, with 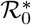 being the basic reproduction number in the absence of intervention programs (*P* = 0). This result reflects the effect of intervention programs to reduce malaria transmission near the DFE.

### B.2 Sensitivity Analysis of the Reduced Model

We conducted a sensitivity analysis on the *ℛ*_0_ for the reduced model to obtain qualitative information on how the model parameters affect ℛ_0_ by employing the normalised forward index, *ζ*, of ℛ_0_ for a parameter *k*, [55] as

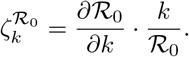

To conduct a thorough analysis of the latency rates in *E*_*H*_ we reparameterised by defining *θ*_*I*_ = *θ · ρ* and *θ*_*A*_ = *θ ·* (1 −*ρ*), where *θ* is the rate of movement from the *E*_*H*_ compartment into the *I*_*H*_ or the *A*_*H*_ compartment, while *ρ* is the probability of becoming clinically infectious after the latent stage in *E*_*H*_ thus moving into *I*_*H*_. We compare the sensitivity index of the parameters on the actual ℛ_0_ (Equation (B.1)) and 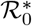 (Equation (12)), and observe that the normalised forward index, *ζ*, of both ℛ_0_ and 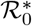 is the same for all parameters except *b, c, η* and *κ* as 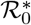 is formulated in the absence of intervention strategies. The results obtained are given in Table 5.

Based on Table 5, we deduce that parameters such as the mosquito biting rate (*δ*), infection success rates (*ψ*_*H*_, *ψ*_*M*_) and the decay rate of intervention programs (*κ*) that have positive indices contribute more to the initial spread of malaria (that is, as the parameter increases, ℛ_0_ increases). Meanwhile, the treatment rate of infectious persons (*τ*), interventions recruitment/funding rate (*η*), and mosquito death rate (*µ*_*M*_) with negative indices reduce ℛ_0_. There are other parameters such as the probability of clinical infectivity rate subsequent to the latent phase (*ρ*) and the disease recovery rate (*γ*) which though suggestive of reducing ℛ_0_, can be manipulated to behave otherwise by altering parameters such as the relative infectiousness of asymptomatic humans (*χ*).

#### B.2.1 Results from sensitivity analysis

Parameter sensitivity findings in Table 5 employing the normalised forward index approach are substantiated in Figure 8.

**Figure 8.**
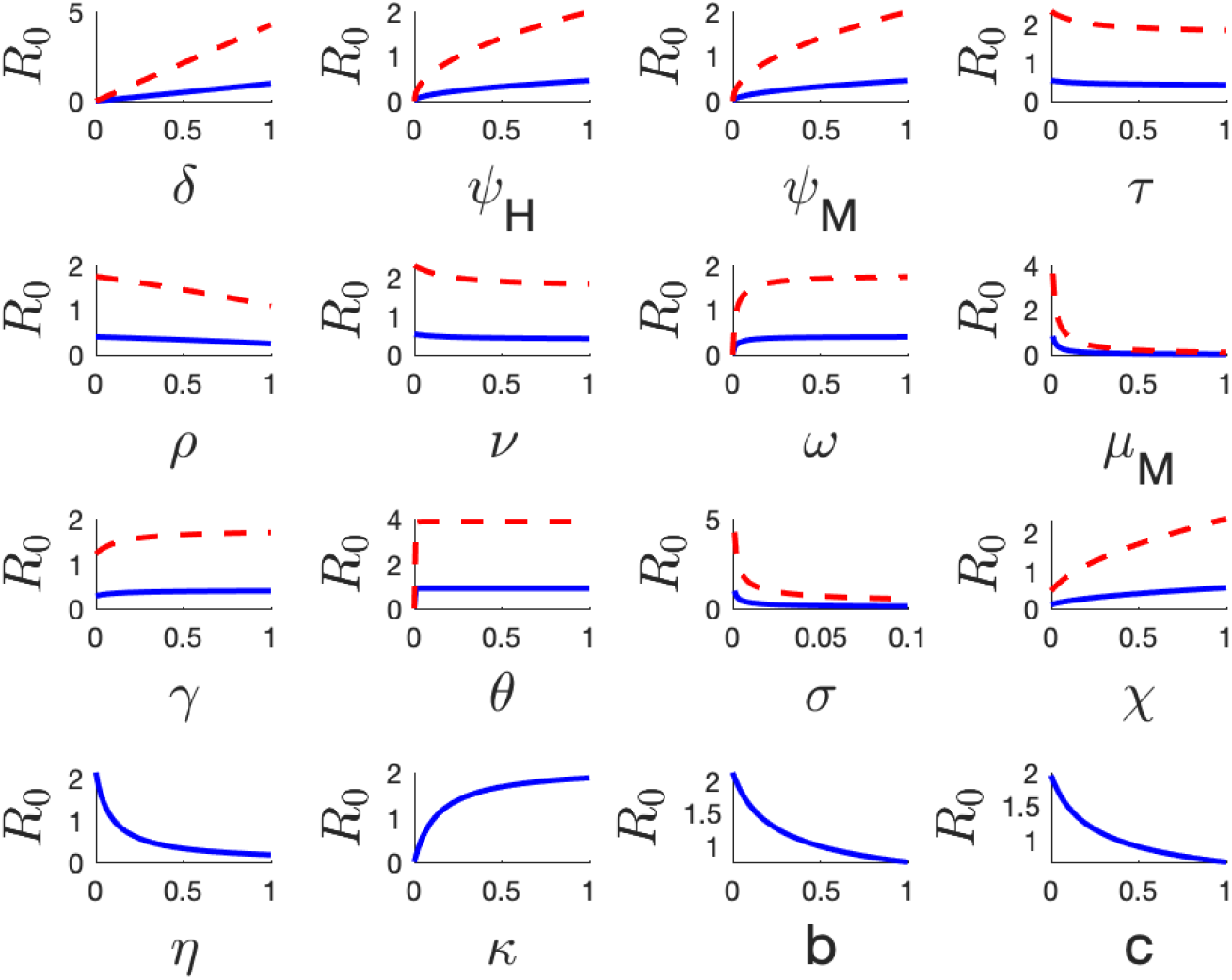
Sensitivity plots resulting from a quantitative local sensitivity analysis conducted on the reduced model. The red dashed curves represent the sensitivity plots of the parameter on 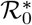 in the absence of interventions while the blue solid lines are obtained from sensitivity analysis on ℛ_0_ with interventions.

**Figure 9.**
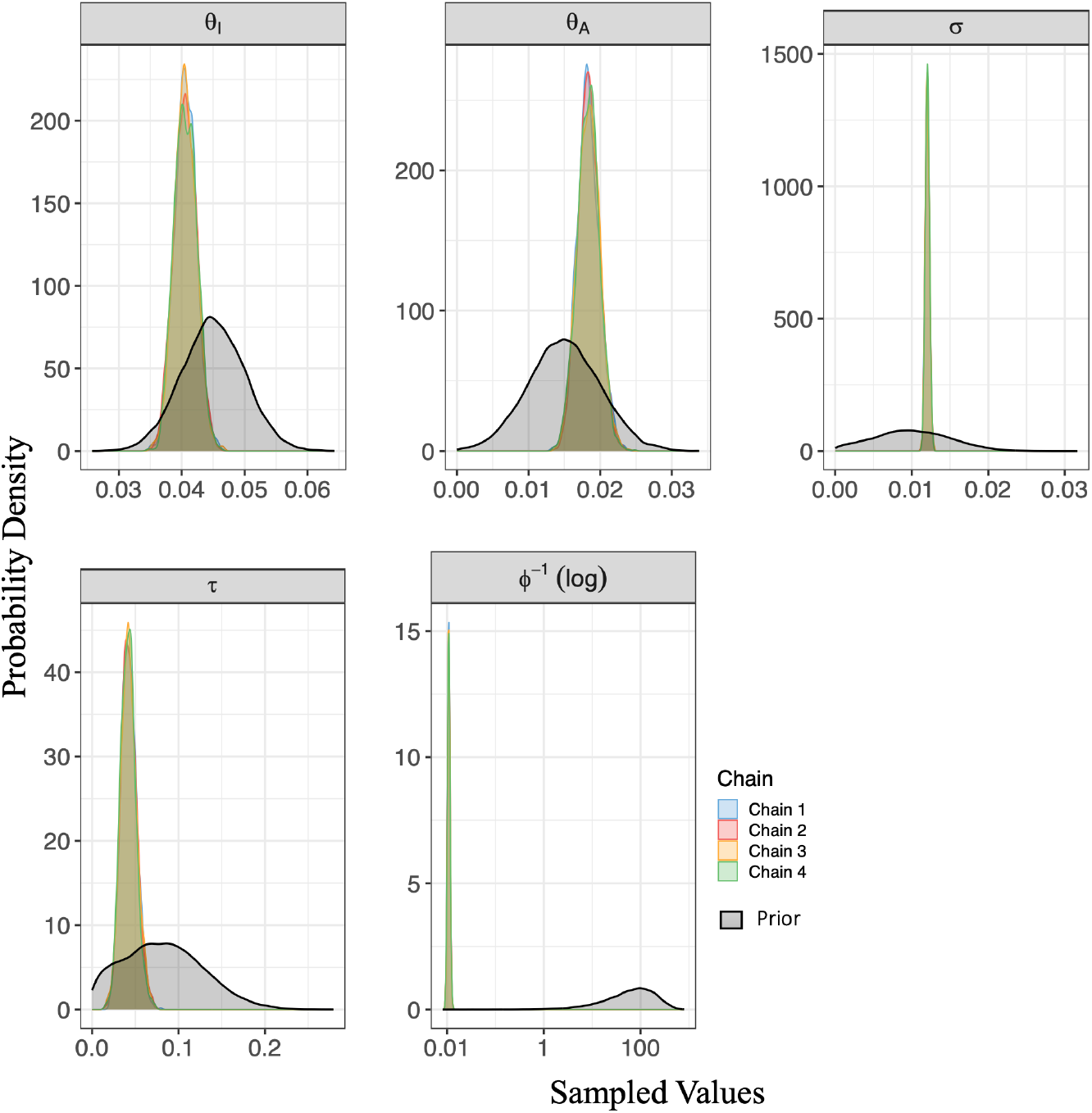
Marginal posterior density and prior distribution plots for the estimated reduced model parameters and the inverse dispersion parameter. The posterior densities confirm coherence among the four Markov chains for the model-to-model inference under a low transmission scenario with 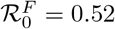 and 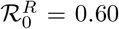 (using the mean estimates of *θ*_*I*_, *θ*_*A*_, *σ* and *τ* from Table 3). Prior distributions are overlaid in black for comparison. The inverse dispersion parameter *ϕ*^−1^ which follows an exponential prior, is presented on a logarithmic scale to accommodate its wide range.

**Figure 10.**
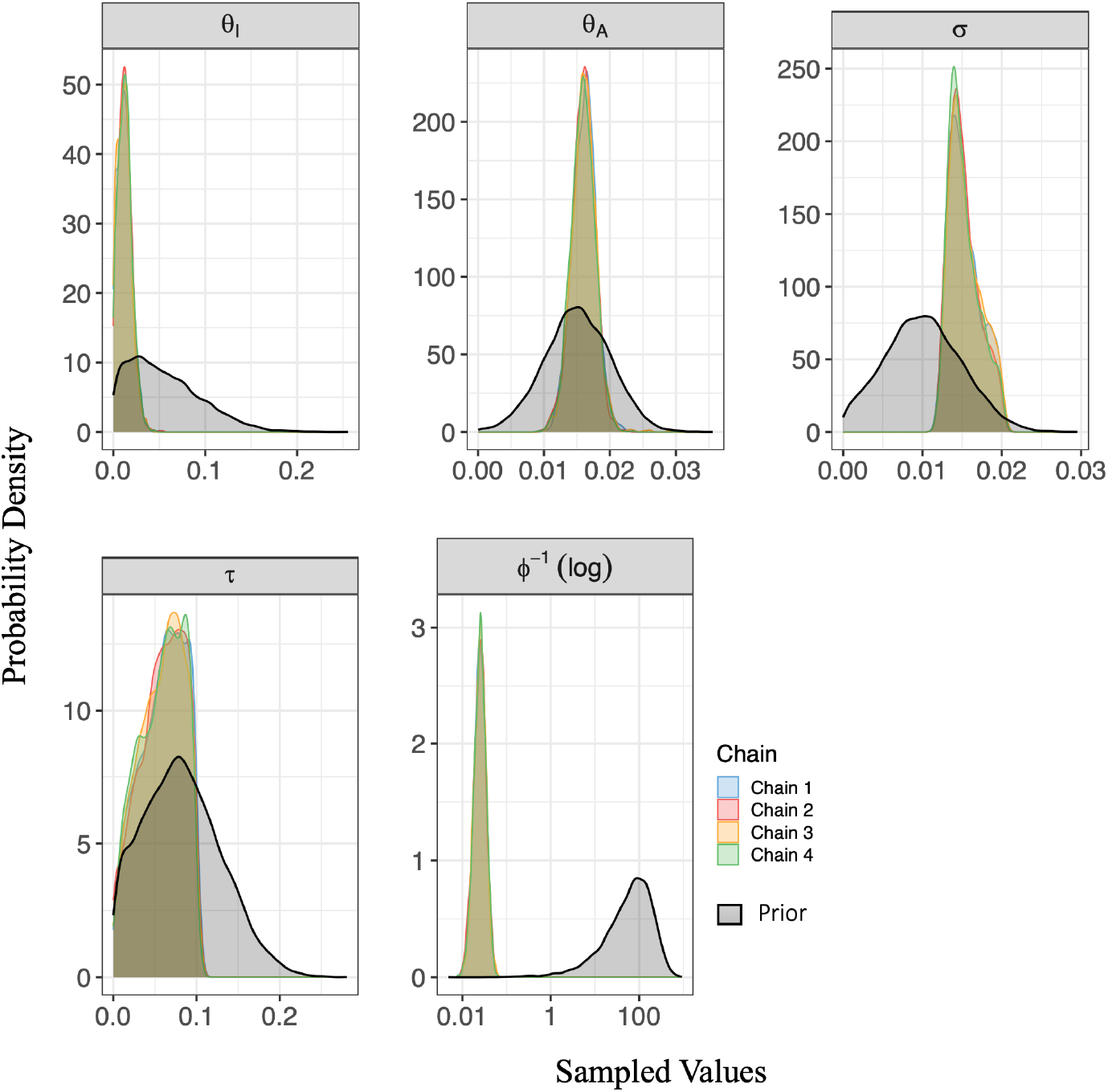
Marginal posterior density and prior distribution plots for the estimated reduced model parameters and the inverse dispersion parameter, confirming coherence amongst the four Markov chains and comparing the posterior distributions against the priors. This result was obtained from the low malaria transmission model-to-model fitting analysis done utilising simulated data from the *A*_*H*_ class of the full model with 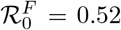 and 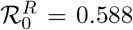 (using the mean estimates from Table 4). Prior distributions are overlaid in black for comparison. The inverse dispersion parameter *ϕ*^−1^ is presented on a logarithmic scale to accommodate its wide range.

Figure 8 illustrates how changes in the reduced model parameters affect the ℛ_0_ of the reduced model. The general observations of Figure 8 confirm our qualitative results in Table 5. In the case of the probability of clinical infectivity after the latent phase, denoted as *ρ*, it is observed that *ρ* has a negative effect on the basic reproduction number. This negative effect occurs, as long as the inequality 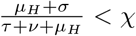 holds, which is the case based on the parameter values in Table 1. Additionally, Figure 8 clearly displays the difference in impact on ℛ_0_ and 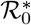 at the baseline parameters, indicating that the absence of intervention programs leads to an increase in the basic reproduction number, which supports the findings from Equation (16).

### B.3 Practical identifiability analysis inference tables

Table 6 summarises the posterior distribution of 13 parameters from the reduced model and the inverse dispersion parameter (*ϕ*^−1^), generated using STAN. Likewise, Table 7 summarises the inference generated for 14 parameters from the full model and the inverse dispersion parameter. We can consider both tables reliable since the Rhat values are close to 1 for all parameters and the effective sample size (n eff) is large, indicating that the Markov chains effectively explored the parameter space [50].

## C Model-to-Model Fitting

### C.1 Bayesian model-to-model fitting

#### C.1.1 Marginal posterior density plots and prior distributions from the low transmission scenario

Building on our results for the low malaria transmission situation (ℛ_0_ *<* 1) explored in Section 3.2, we now examine a high malaria transmission case. We consider the two scenarios of fitting the reduced model to data from all compartments and the *A*_*H*_ class as considered in Section 3.2 and set *δ* = 2 while keeping all other parameters as per Table 1. With 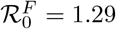, we generate noisy simulation data from the full model, utilising the same initial conditions from Section 3.2.

##### Fitting using data from all full model compartments (high transmission scenario)

Supposing that all compartments of the full model are observed in an endemic population at 5-day intervals over a four-year period. We represent the full system *F* (*t*) with a negative binomial distribution of the reduced system *R*(*t*) while accommodating over-dispersion by incorporating the parameter *ϕ*, as done in Section 3.2. The negative binomial formulation used to generate the simulation data is of the form

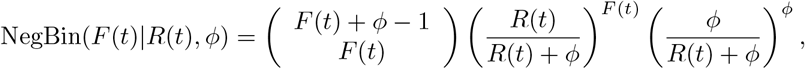

at every time point t. We fit the reduced model to the observed data (full model) to obtain a posterior distribution for the model parameters. The prior distribution of the model parameters, *θ* = (*θ*_*I*_, *θ*_*A*_, *σ, τ, ϕ*^−1^) was maintained as defined in Section 3.2.

The results obtained for this scenario are highlighted in Table 8, Figures 11, 12 and 13. Using the mean estimates for the reduced model in Table 8, the reproduction number of the reduced model was 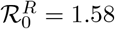, while the data was generated with the full model at 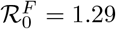.

**Table 8:**
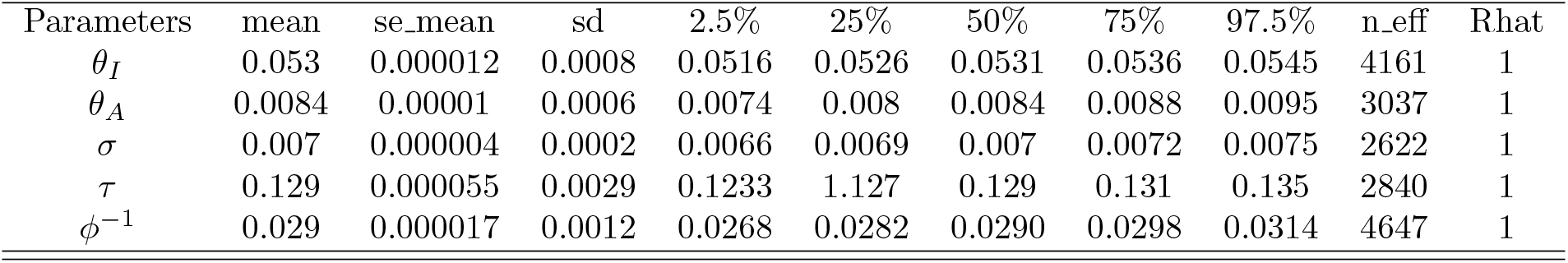
Summary of Bayesian parameter inference obtained from the model-to-model fitting considering a combined dataset from all compartments of the full model assuming a high transmission situation with 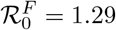. The table provides posterior estimates for the reduced model’s parameters *θ*_*I*_, *θ*_*A*_, *σ* and *τ*, as well as the inverse dispersion parameter *ϕ*^−1^.

| Parameters | mean | se_mean | sd | 2.5% | 25% | 50% | 75% | 97.5% | n_eff | Rhat |
| --- | --- | --- | --- | --- | --- | --- | --- | --- | --- | --- |
| $\theta_I$ | 0.053 | 0.000012 | 0.0008 | 0.0516 | 0.0526 | 0.0531 | 0.0536 | 0.0545 | 4161 | 1 |
| $\theta_A$ | 0.0084 | 0.00001 | 0.0006 | 0.0074 | 0.008 | 0.0084 | 0.0088 | 0.0095 | 3037 | 1 |
| $\sigma$ | 0.007 | 0.000004 | 0.0002 | 0.0066 | 0.0069 | 0.007 | 0.0072 | 0.0075 | 2622 | 1 |
| $\tau$ | 0.129 | 0.000055 | 0.0029 | 0.1233 | 0.127 | 0.129 | 0.131 | 0.135 | 2840 | 1 |
| $\phi^{-1}$ | 0.029 | 0.000017 | 0.0012 | 0.0268 | 0.0282 | 0.0290 | 0.0298 | 0.0314 | 4647 | 1 |

**Figure 11.**
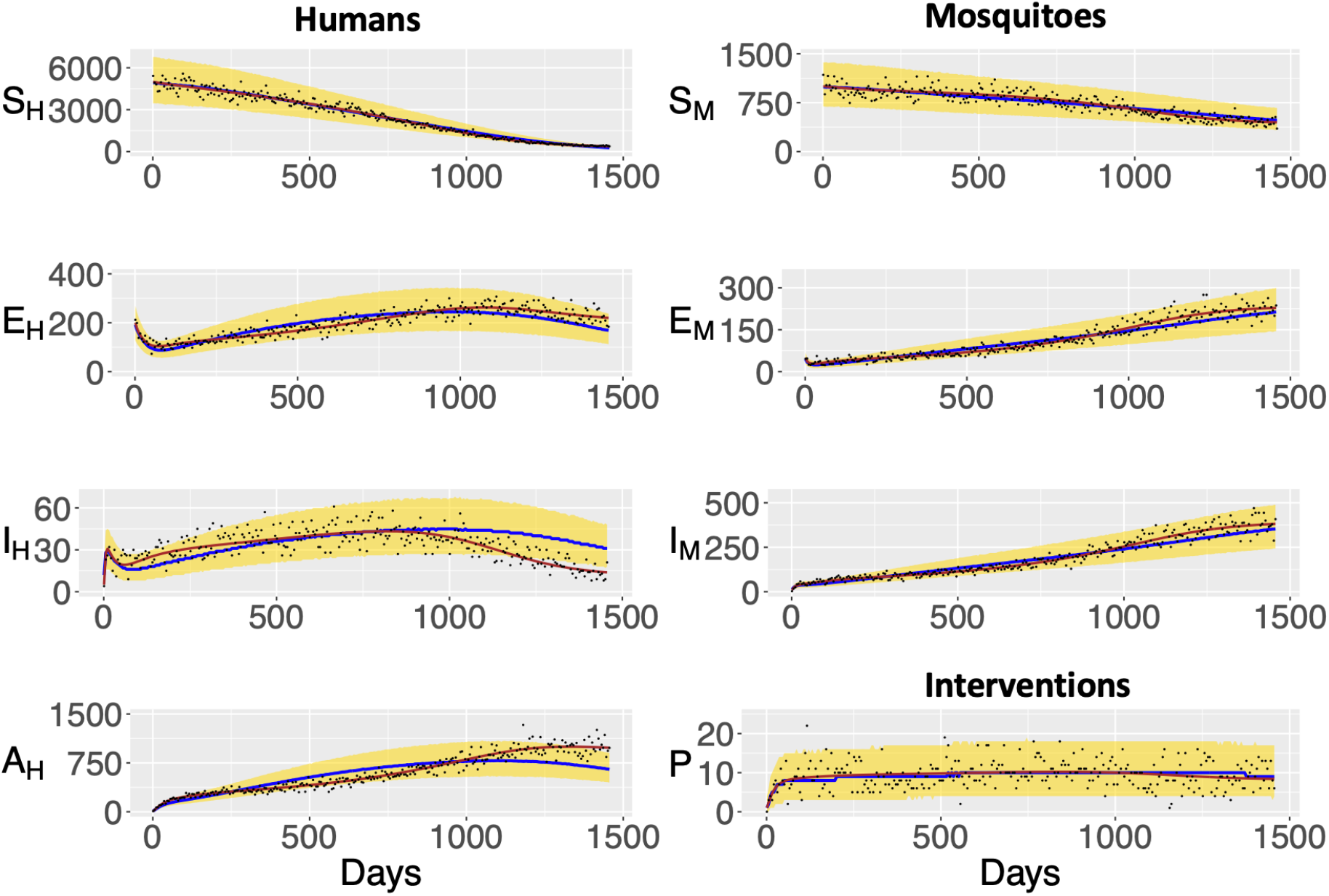
Posterior predictive check of the reduced model compartments. The blue solid line is the median line and the yellow region, the 95% prediction interval of the reduced model. The black points represent the noisy simulated data points generated from the combined full model compartments whereas the brown solid line represents the actual simulation data generated from the full model compartments without noise. These results were obtained from a model-to-model fitting analysis done utilising simulated data from the full model with 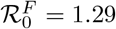 and 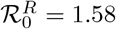 (using the mean estimates from Table 8).

**Figure 12.**
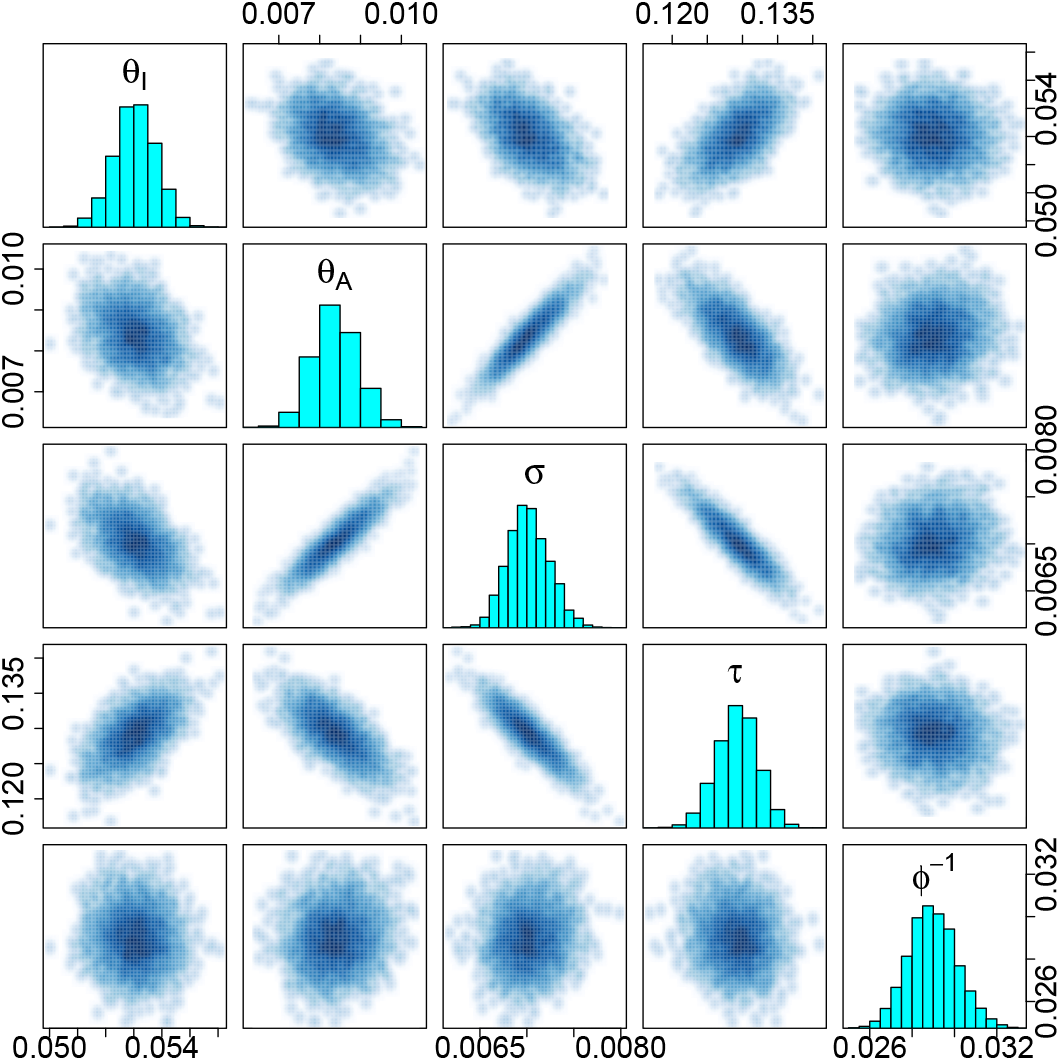
Pairwise plots of the estimated reduced model parameters; *θ*_*I*_, *θ*_*A*_, *σ* and *τ*, and the inverse dispersion parameter *ϕ*^−1^ from the inference generated from the fitting analysis observing the compartments the full model and assuming a high transmission scenario with 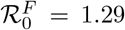 and 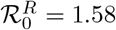 (using the mean estimates from Table 8).

**Figure 13.**
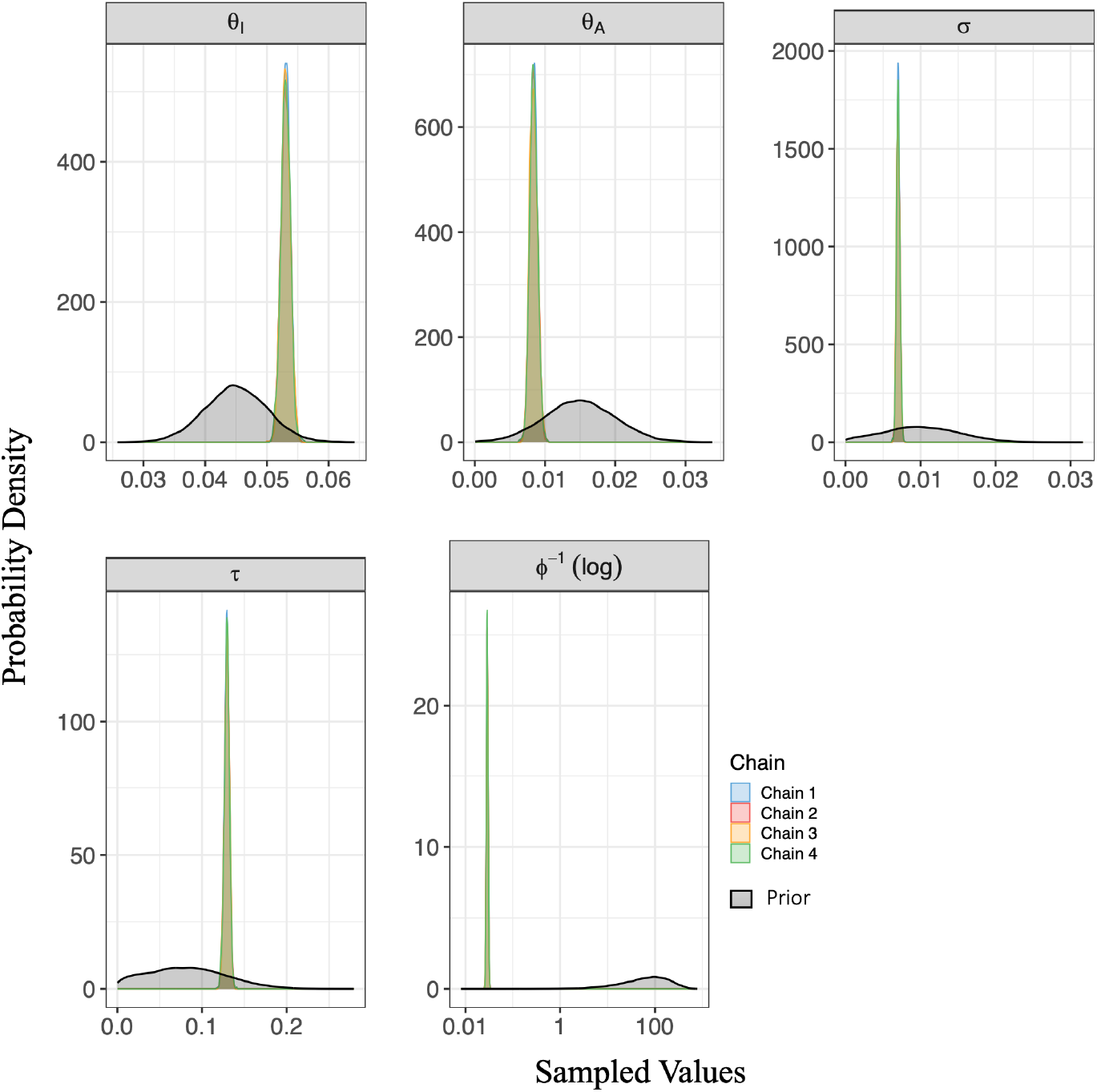
Marginal posterior density and prior distribution plots for the estimated reduced model parameters and the inverse dispersion parameter. The posterior densities confirm coherence among the four Markov chains. This result was obtained from a model-to-model inference assuming a high transmission scenario with 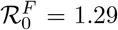 and 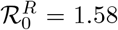 (per the mean estimates from Table 8). Prior distributions are overlaid in black for comparison. The immunity waning rate *σ* and inverse dispersion *ϕ*^−1^ parameters are presented on logarithmic scales to accommodate their wide range.

From Figure 11, we observe that a significant number of the noisy datapoints of the full model (black dots), median prediction line (blue curve), and the actual simulation data without noise (brown curve) lie within the 95% prediction interval (yellow region) for the mosquito compartments and interventions class. For the human compartments, we observe the same dynamics for the susceptible *S*_*H*_, exposed *E*_*H*_ and asymptomatic infectious *A*_*H*_ compartments during the entire four-year period. The same is noticed in the first three years for the clinically infectious human class *I*_*H*_. However, within the fourth year, the 95% prediction interval of the clinically infectious class *I*_*H*_ does not capture several noisy simulated data points as well as the brown curve which is the simulation data without noise. Moreover, the reduced model predicts a higher number of clinically infectious cases than observed from the full model, as the median prediction line (blue curve) and yellow region are higher than the brown line (actual simulation data from the full model) and the black dots, (noise-incorporated simulation data from the full model), in the *I*_*H*_ plot of Figure 11. This behaviour could result from the estimation of a higher *θ*_*I*_ value, see Table 8 as compared to values observed in Tables 1 and 3 and subsequently a higher estimated reproduction number of the reduced model (given the mean estimates from Table 8) than the full model as 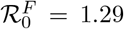 and 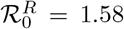. While this behaviour was observed for the clinically infectious human class in the fourth year, it is worth noting that the difference between the estimated number of clinically infectious cases of the reduced model and the observed number of clinically infectious cases of the full model is less than 20 clinical infectious humans per 5 days within the fourth year period. Thus while the data points (black dots-noisy simulation data from the full model) are not fully captured in the fourth year, the predicted number of clinically infectious cases of the reduced model is close to the observed number of clinically infectious cases of the full model.

We observe from Table 8 a high level of agreement among the four Markov chains, which is further confirmed by observing Figure 13. From Figure 13 the posterior densities appear substantially more concentrated than the prior distributions across all parameters, indicating increased precision and strong information from the data. Additionally, the effective sample size of the estimated parameters indicates that the Markov chains effectively explored the parameter space [50]. From Figure 12, we observe positive correlations between *σ* and *θ*_*A*_, and *θ*_*I*_ and *τ*, negative correlations between *σ* and *τ*, *θ*_*I*_ and *θ*_*A*_, and *θ*_*I*_ and *σ*. These posterior behaviours are expected given the combined nature of the immunity pathways (considered separate in the full model) in the reduced model, see Figure 2. For instance, the direct correlation between *θ*_*I*_ and *τ* implies that an increase or decrease in the symptomatic latency rate directly influences the treatment rate, indicating that an increasing number of symptomatic/clinically infectious individuals results in an increased treatment of the symptomatic cases, see Figure 2.

#### C.1.3 Fitting results using data from asymptomatic infectious humans *A*_*H*_ only (high transmission scenario)

We proceed to consider a more realistic scenario of the high transmission case by only observing the asymptomatic infectious class data generated from the full model. The results obtained for this scenario are highlighted in Table 9, Figures 14, 15 and 16. Given the mean estimates for the reduced model in Table 9, the reproduction number of the reduced model was 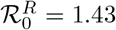, while the data was generated with the full model at 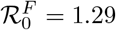.

**Table 9:** Summary table of Bayesian parameter inference obtained from the model-to-model fitting considering data from only the asymptomatic class of the full model, assuming a high transmission situation with 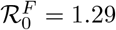. The table provides posterior estimates for the reduced model’s parameters *θ*_*I*_, *θ*_*A*_, *σ* and *τ*, as well as the inverse dispersion parameter *ϕ*^−1^.

| Parameters | mean | se.mean | sd | 2.5% | 25% | 50% | 75% | 97.5% | n.eff | Rhat |
| --- | --- | --- | --- | --- | --- | --- | --- | --- | --- | --- |
| $\theta_I$ | 0.179 | 0.00044 | 0.0297 | 0.125 | 0.158 | 0.178 | 0.198 | 0.242 | 4624 | 1 |
| $\theta_A$ | 0.033 | 0.00008 | 0.0053 | 0.024 | 0.029 | 0.033 | 0.036 | 0.044 | 4053 | 1 |
| $\sigma$ | 0.0068 | 0.000004 | 0.0003 | 0.0062 | 0.0066 | 0.0068 | 0.0070 | 0.0074 | 5800 | 1 |
| $\tau$ | 0.265 | 0.00031 | 0.022 | 0.225 | 0.249 | 0.263 | 0.279 | 0.311 | 5125 | 1 |
| $\phi^{-1}$ | 0.034 | 0.00004 | 0.0031 | 0.028 | 0.032 | 0.034 | 0.036 | 0.040 | 6379 | 1 |

**Figure 14.**
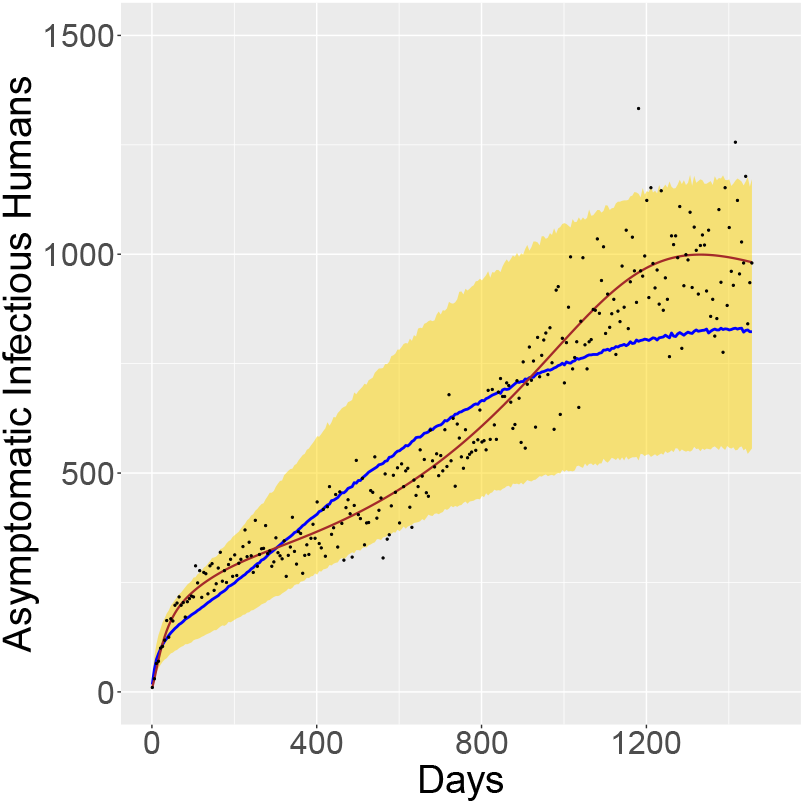
Posterior predictive check of the asymptomatic infectious human class of the reduced model. The blue solid line is the median line and the yellow area, the 95% prediction interval of the reduced model’s asymptomatic infections. The black points represent the noise-incorporated simulated data points generated from the full model whereas the brown solid line represents the smooth simulation data generated from the full model without noise. This result was obtained from a model-to-model fitting analysis done assuming a high transmission scenario with 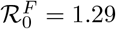 and 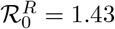 (using the mean estimates from Table 9).

**Figure 15.**
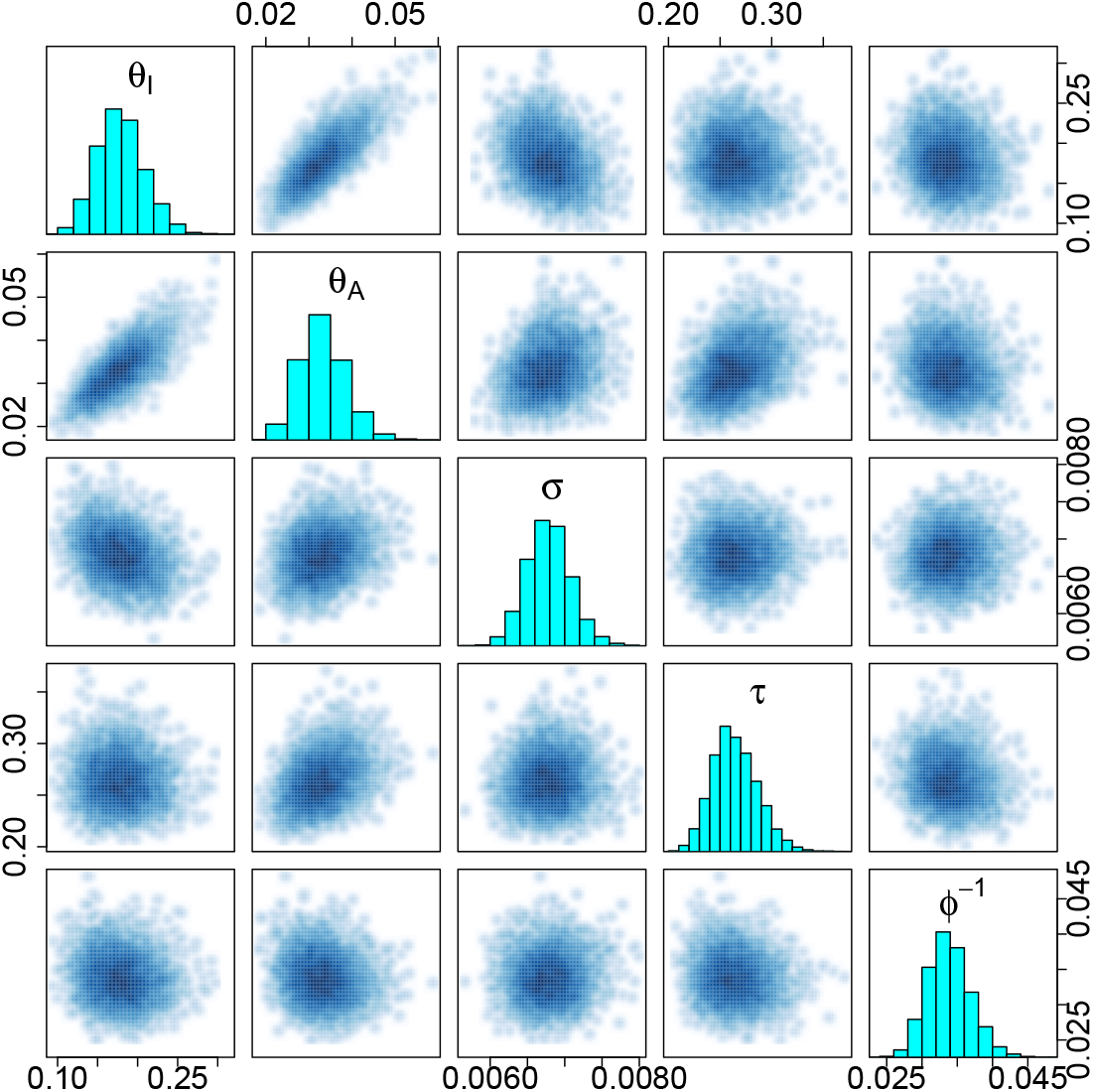
Pairwise plots of the estimated reduced model parameters; *θ*_*I*_, *θ*_*A*_, *σ* and *τ*, and the inverse dispersion parameter *ϕ*^−1^ from the inference generated from the fitting analysis observing only the *A*_*H*_ class of the full model, assuming a high transmission scenario with 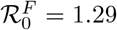 and 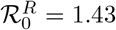 (using the mean estimates from Table 9).

**Figure 16.**
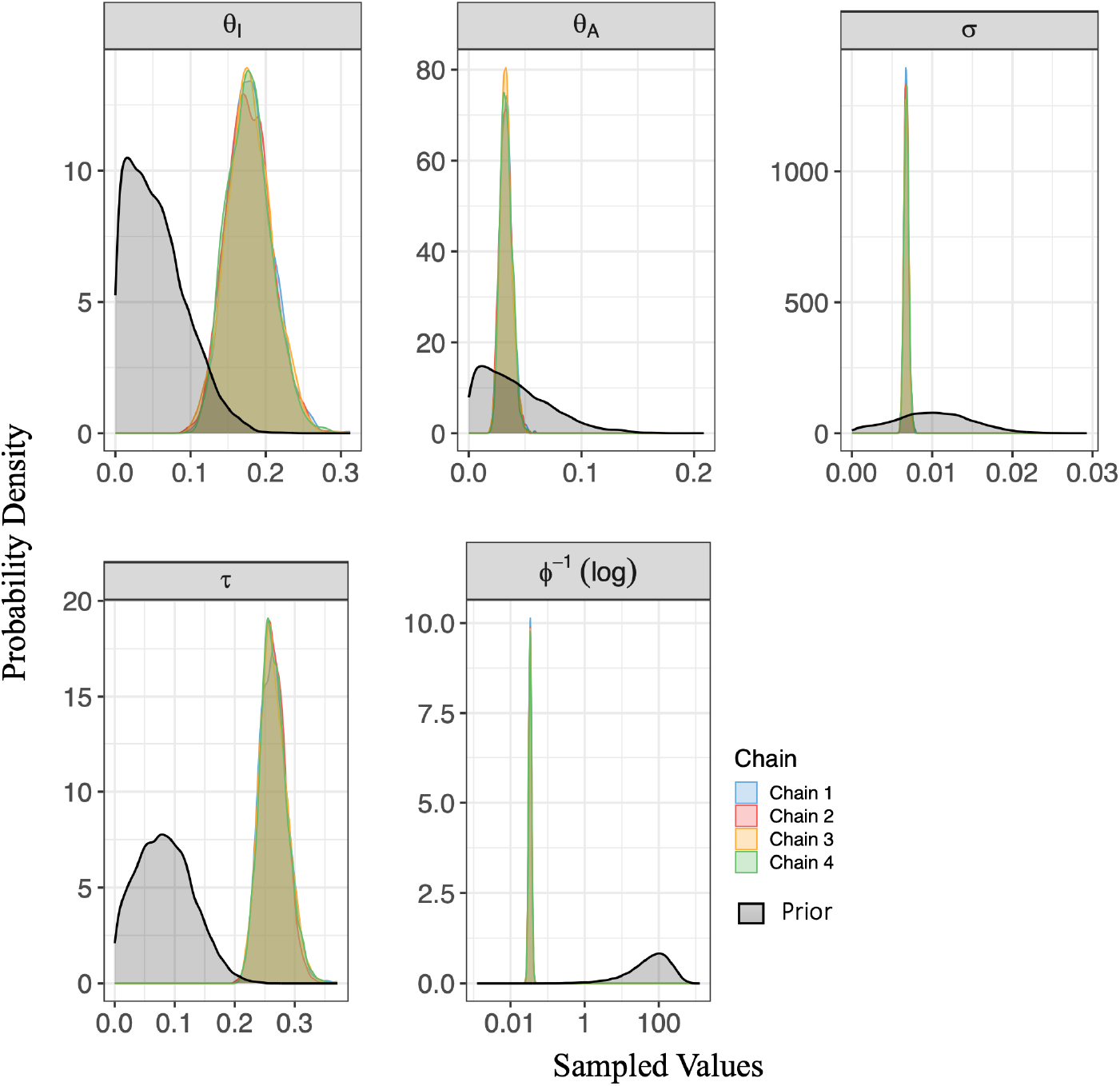
Marginal posterior density and prior distribution plots for the estimated reduced model parameters and the inverse dispersion parameter, confirming coherence amongst the four Markov chains and comparing the posterior distributions against the priors. This result was obtained from the high malaria transmission scenario model-to-model fitting analysis done utilising simulated data from the *A*_*H*_ class of the full model with 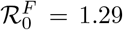 and 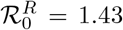 (using the mean estimates from Table 9). Prior distributions are overlaid in black for comparison. The inverse dispersion *ϕ*^−1^ parameter is presented on a logarithmic scale to accommodate its wide range.

Figure 14 demonstrates a good fit of the reduced model’s asymptomatic infectious class to the actual simulated data (brown solid line) and noisy data (black points) from the full model’s asymptomatic infectious class as the posterior predictive check (Figure 14) shows that the noisy data and the smooth simulated data lie within the 95% prediction interval (yellow shade). Results from the inference table, Table 9 estimates the reduced model parameters; *θ*_*I*_, *θ*_*A*_, *σ* and *τ* as well as the inverse dispersion parameter. The convergence of the four Markov chains is confirmed by the Rhat values and the effective sample size in Table 9 as well as the marginal posterior density plots in Figure 16, which also indicates the efficient exploration of the parameter space [50]. In comparing the inference tables, Tables 8 and 9, we observe that while there is agreement among the four chains considered in two high transmission fitting scenarios, the estimated parameters of the reduced model have comparably lower standard mean errors in the scenario where all classes of the full model are observed than in the scenario where only the asymptomatic class, *A*_*H*_ of the full model is observed. This indicates that more reliable parameter estimations were obtained in the scenario in which all classes of the full model are observed. From the correlation plots in Figure 15, a clear positive correlation is observed between *θ*_*I*_ and *θ*_*A*_, indicating that the two parameters are not fully identifiable from each other since they show significant correlation.

The other parameters; *σ, τ* and *ϕ*^−1^ appear relatively well-identifiable as their joint distributions form roughly circular clouds, suggesting independence. The positive correlation between *θ*_*I*_ and *θ*_*A*_ suggests that an increase or decrease in the symptomatic latency rate directly influences the asymptomatic latency rate, which would be the case in the long run, see Figure 2, given a high treatment rate, see the estimated treatment rate parameter, *τ* value in Table 9 in comparison with values in Tables 8, 3, and 4.

Overall, our findings suggest that the reduced model effectively approximates Korsah et al.’s model and exhibits similar transmission characteristics of the model while offering improved amenability for further studies focused on exploring the impact of malaria interventions and the transmission dynamics of specific endemic areas.

